# Post-acute health care costs following SARS-CoV-2 infection: A retrospective cohort study of among 531,182 matched adults

**DOI:** 10.1101/2023.08.02.23293563

**Authors:** Candace D. McNaughton, Peter C. Austin, Zhiyin Li, Atul Sivaswamy, Jiming Fang, Husam Abdel-Qadir, Jacob A. Udell, Walter Wodchis, Douglas S. Lee, Ivona Mostarac, Clare L. Atzema

## Abstract

Post-acute health care costs following SARS-CoV-2 infection are not known. Beginning 56 days following SARS-CoV-2 polymerase chain reaction (PCR) testing, we compared person-specific total and component health care costs across their distribution for the following year (test-positive versus test-negative, matched people; January 1, 2020-March 31, 2021). For 531,182 individuals, mean person-specific total health care costs were $513.83 (95% CI $387.37-$638.40) higher for test-positive females and $459.10 (95% CI $304.60-$615.32) higher for test-positive males, or >10% increase in mean per-capita costs, driven by hospitalization, long-term care, and complex continuing care costs. At the 99^th^ percentile of each subgroup, person-specific health care costs were $12,533.00 (95% CI $9,008.50-$16,473.00) higher for test-positive females and $14,604.00 (95% CI $9,565.50-$19,506.50) for test-positive males, driven by hospitalization, specialist (males), and homecare costs (females). Cancer costs were lower. Six-month and 1-year costs differences were similar. These findings can inform planning for post-acute SARS-CoV-2 health care costs.

## Introduction/Background

It is difficult to overstate the importance of the COVID-19 pandemic as a new public health threat, with more than a total of 6.8 million deaths worldwide and at least half of Canadians infected as of August 2022.^1–3^ In the relatively new context of widespread, on-going transmission of this airborne pathogen, there is growing recognition of the considerable health burden and costs beyond the acute phase of COVID-19.^4^ There is growing evidence that infection caused by the airborne SARS-CoV-2 virus has longer-term health effects such as increased risk of metabolic derangements, auto-immune conditions, and cardiovascular disease, as well as evidence that post-acute complications can occur across the spectrums of age and severity of the acute infection.^5–10^ COVID-19 and its downstream costs are among the most pressing challenges to the long-term health of Canadians and sustainability of the Canadian health care system.^11^

Health care use following SARS-CoV-2 infection is higher than in otherwise similar, uninfected individuals, even months following infection and among those who were not hospitalized acutely, and the distribution of health care use increase is right skewed, with a small proportion of individuals experiencing markedly greater health care use.^12, 13^ However, differences in direct health care costs following infection are not well understood and, in light of the sheer number of on-going infections, are necessary to understand in order to better anticipate and plan for the magnitude, duration, and types of potential increases in health care costs.

We therefore compared health care costs of community-dwelling adults starting ≥56 days following a positive SARS-CoV-2 polymerase chain reaction (PCR) to matched individuals who tested negative for SARS-CoV-2.

## Methods

### Study Design

This retrospective cohort was constructed using population-level administrative and health care data linked and held at ICES, formerly known as the Institute for Clinical Evaluative Sciences. ICES is a non-profit research institute funded by, but independent of, the Ontario Ministry of Health and the Ontario Ministry of Long-Term Care to evaluate health care delivery and outcomes to inform health care policies. In accordance with Ontario’s Personal Health Information Protection Act (section 45) permitting collection and analysis of health care and demographic data for health system evaluation, individual consent was not obtained.^14^

### Cohort creation

All community-dwelling adults (≥18 years of age) in Ontario, Canada with sufficient demographic information for data linkage (valid date of birth, sex, and death information) who underwent PCR testing for SARS-CoV-2 between January 1, 2020 and March 31, 2021 and were alive eight weeks (56 days) after the date of PCR test were included. During this time, SARS-CoV-2 PCR tests were generally widely available and free-of-charge, and testing was performed entirely within the health care system of Ontario, which is pays for all necessary physician and hospital services for >98% of the 14.2 million residents of Ontario. Datasets used for cohort construction and variable definitions are listed in Supplemental Tables E1 and E2, respectively.

### Exposure definition

Individuals were categorized as test-positive as of the date of their first positive outpatient SARS-CoV-2 PCR test. Individuals were categorized as test-negative if all PCR results during the study time were negative. PCR results reported as pending or indeterminate (<0.02%) were excluded. The index date was defined as the date of first positive outpatient SARS-CoV-2 PCR test or, for individuals with only negative PCR test(s), the last test date. Symptoms of acute SARSC-CoV-2 infection generally resolve within eight weeks,^15–18^ follow-up to assess post-acute health care costs began eight weeks (56 days) after the index date and ended on May 26, 2022 or death, whichever occurred first.

### Matching

Individuals with a positive SARS-CoV-2 PCR test result were matched to those with only negative test results by sex/pregnancy status, pandemic period (2-week intervals), public health unit, vaccination status, and a propensity score computed from a large set of demographic, clinical, and health services factors, including component health care costs in year prior to the index PCR date, age, baseline socio-demographics and clinical characteristics, neighborhood level socioeconomic indices, and COVID vaccination status (Table E2).^19^ Subjects were matched on the logit of the propensity score using a caliper width equal to 0.05 times the standard deviation of the propensity score.^20^ A standardized difference <0.1 for comparing baseline characteristics between exposure groups was considered as indicative of good balance between exposure groups.^19, 21^

### Outcome definitions

The primary outcome was the person-specific cost of health care over the first year of follow-up, i.e., the amount of money per-person spent by the Ontario Ministries of Health and Long Term Care, which pays for all medical care in the universal health care system. Costs for physician and hospital services are standard across Ontario and include the following 11 mutually exclusive component costs: hospital-based care costs; emergency department visit costs; outpatient (primary care, specialist, cancer care) costs; outpatient medication costs (for people aged ≥65 years); outpatient laboratory costs; rehabilitation costs; mental health hospitalization costs; complex and continuing care costs; homecare costs; and long-term care costs. Because types of health care vary by health care system, descriptions of component health care costs are included in Supplemental Table E1. Observed total and component health care costs in the period starting 56 days after the index PCR test date were summed and divided by the number of days of follow-up (i.e., exposure time), and then multiplied by 365 days to calculate annual per-person costs. For patients hospitalized at the start of follow-up, hospital-based costs were prorated so that only hospital-based costs that occurred during follow-up were included.

Cost differences were computed for test-positive versus test-negative individuals at the mean, median, 95^th^ percentile, and 99^th^ percentiles of health care costs. Note that mean component costs are additive, but this is not the case for median, 95^th^ percentile, or 99^th^ percentile component costs. All costs were standardized to the value of the Canadian dollar in 2020.

Secondary outcomes included the following dichotomous outcomes for outpatient cancer, rehabilitation, home care, mental health admission, complex continuing care, and long-term care costs, overall and stratified by sex: 1) being a high-cost health care user, defined as > 95^th^ percentile of total health care costs from the matched cohort,^22, 23^ and 2) any reported post-acute health care cost, i.e., evidence of any post-acute health care seeking behaviour or access. We also estimated the risk of entrance to long-term care (i.e., nursing home) during 6-month and 1-year follow-up.

### Statistical Analyses

Baseline characteristics of the unmatched and matched cohorts were reported as means with standard deviations (SD), medians and interquartile ranges (IQR), or frequencies, as appropriate. Total and component annualized health care costs based on 1-year and 6-month follow-up were reported as means (SDs) and medians (Q1, Q3).

Based on prior studies indicating interaction by sex,^13, 24, 25^ we assessed for potential interaction by sex in the matched cohort using a P-value threshold <0.10. This revealed evidence of interaction with some component costs (outpatient medication, outpatient primary care, outpatient laboratory, rehabilitation care, homecare, mental health admissions, and long-term care component costs; Supplemental Table E3). Therefore, analyses were performed for the overall cohort as well as stratified by sex.

To compare health care costs across a right-skewed distribution, person-specific annualized costs for matched test-positive versus negative individuals were compared at the mean (by paired T-test), median, 95^th^ percentile, and 99^th^ percentile (by Wilcoxon signed rank test),^26^ and 95% CI’s were generated by bootstrapping with 2000 bootstrap replicates.^27^ Sensitivity analyses censored at admission to long-term care and six months. Relative risks (RR) were computed for 6-month and 1-year high-cost health care use and any post-acute health care for six component costs (outpatient cancer care, rehabilitation, home care, mental health hospitalization, complex continuing care, and long-term care). The 95% CI RR were estimated using methods appropriate for matched data.^28^ All analyses were performed in SAS v. 9.04.

## Results

Between January 1, 2020 and March 31, 2021 there were >11 million SARS-CoV-2 PCR tests completed for 3,777,451 unique adults. Of the 3,631,040 individuals who met the study inclusion criteria, 268,521 (7.4%) had a positive SARS-CoV-2 PCR test, and 99% of test-positive individuals were successfully matched to a test-negative control. The resulting matched cohort consisted of 531,182 individuals (Supplemental Figure E1).

Demographics, clinical characteristics, and standardized differences comparing the distribution of baseline characteristics between positive and negative individuals for the matched and the unmatched cohorts are reported in Table 1 and Supplemental Table E4, respectively. Compared to the unmatched cohort, the matched cohort was slightly younger and had higher proportion of males, urban, lower income, ethnically diverse neighborhoods with overall slightly lower prevalence of comorbid conditions and lower baseline health care costs. In addition, individuals in the matched cohort were less likely to have received any COVID vaccine dose, and they more often underwent PCR testing during late 2020 or early 2021 compared to the unmatched cohort.

**Table 1:**
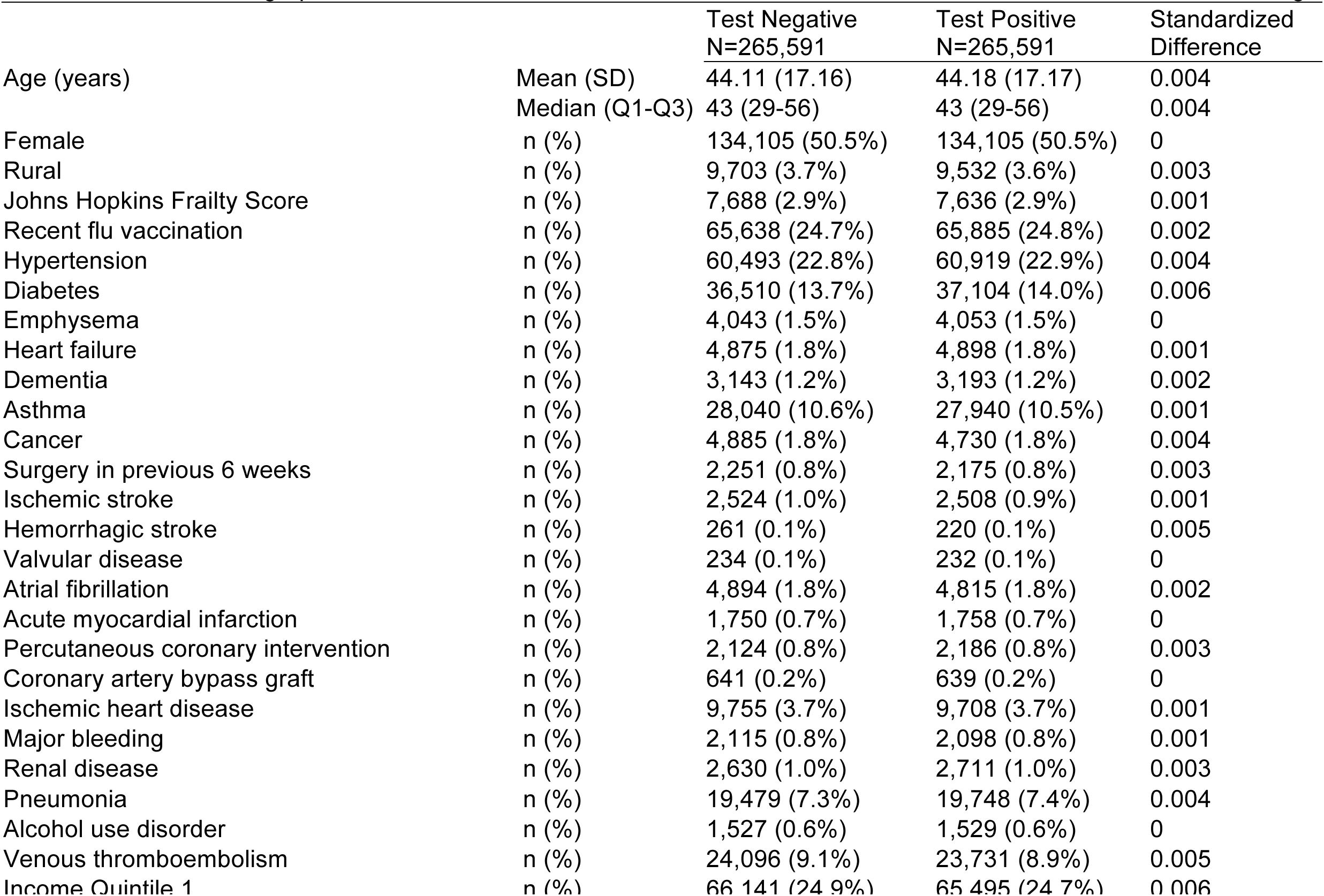
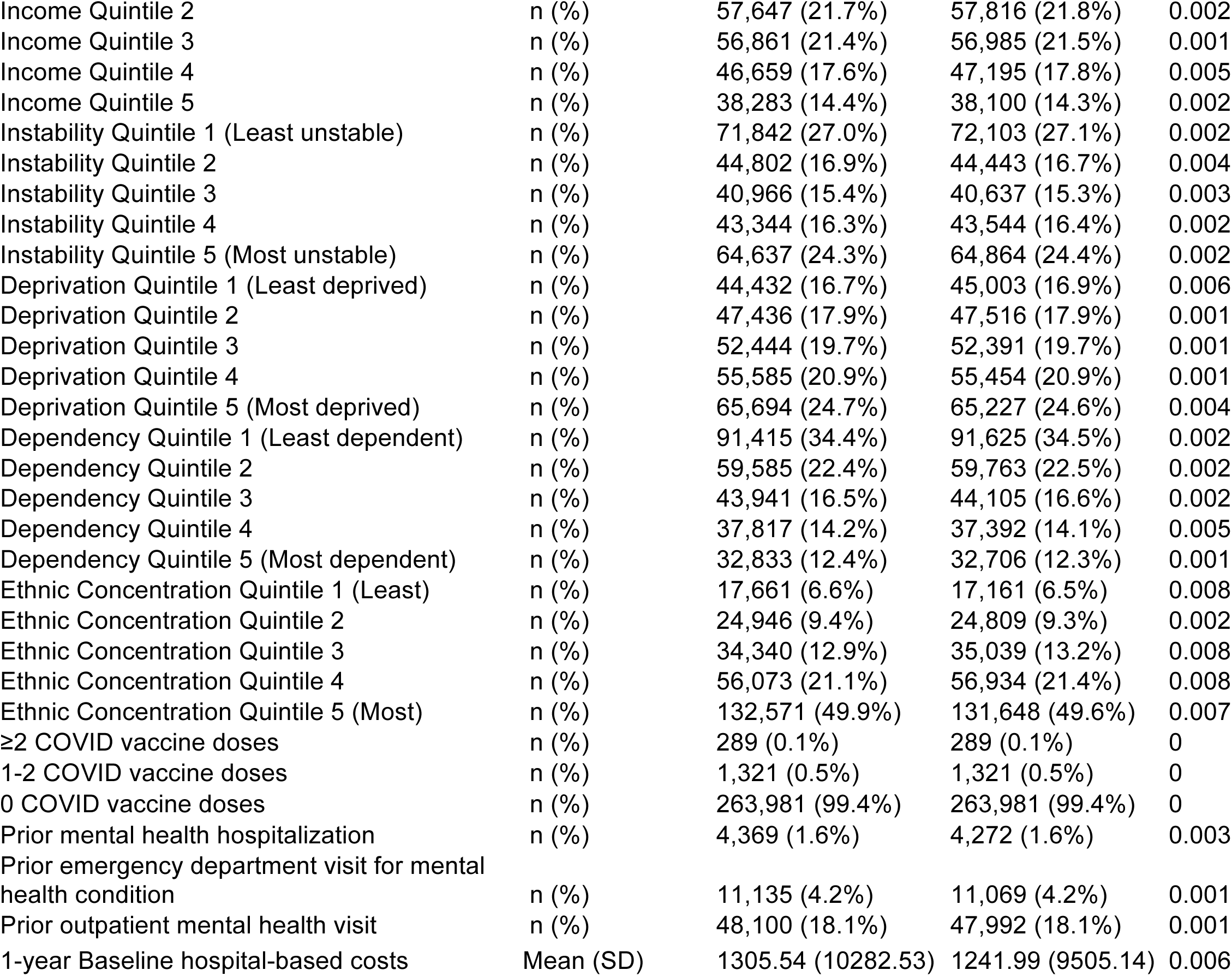
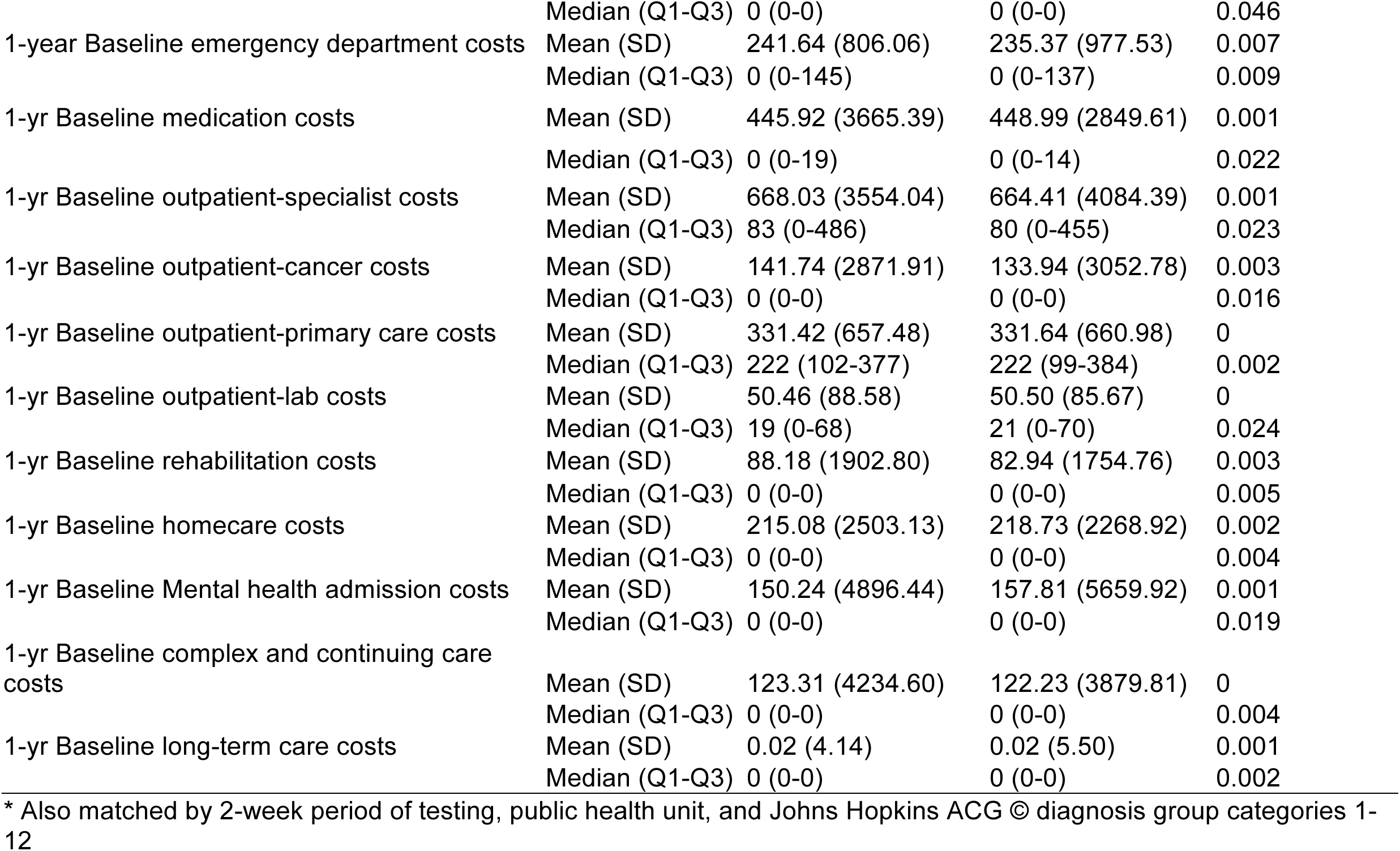
Baseline demographics and clinical characteristics of the matched cohort and characteristics used for matching*.

### Total person-specific annualized health care costs

Differences in annual total and component health care costs per person, overall and stratified by sex, comparing matched test-positive and test-negative people are shown in Figure 1 (comparison of mean annualized costs, range -$100 to +$600) and Figure 2 (comparison of 99^th^ percentile annualized costs, range -$1,000 to +20,000). Comparisons at median and 95^th^ percentiles are found in Supplemental Figures E2 and E3, respectively.

**Figure 1:**
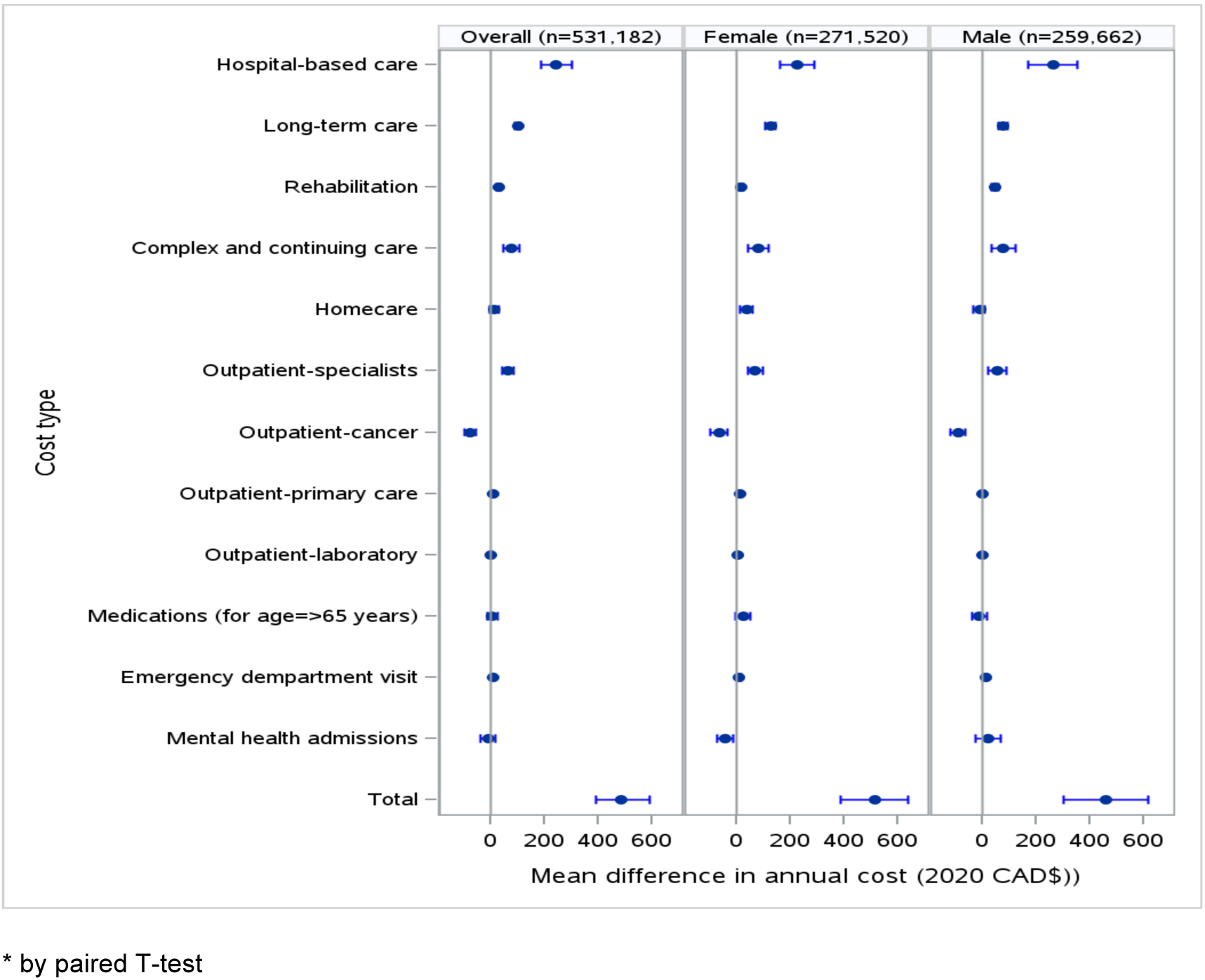
Differences in person-specific annual mean health care costs* for test-positive versus test-negative matched individuals, overall and by sex (95% confidence intervals in 2020 $CAD; starting 56 days after polymerase chain reaction test). Note range -$200 to +$600.

**Figure 2:**
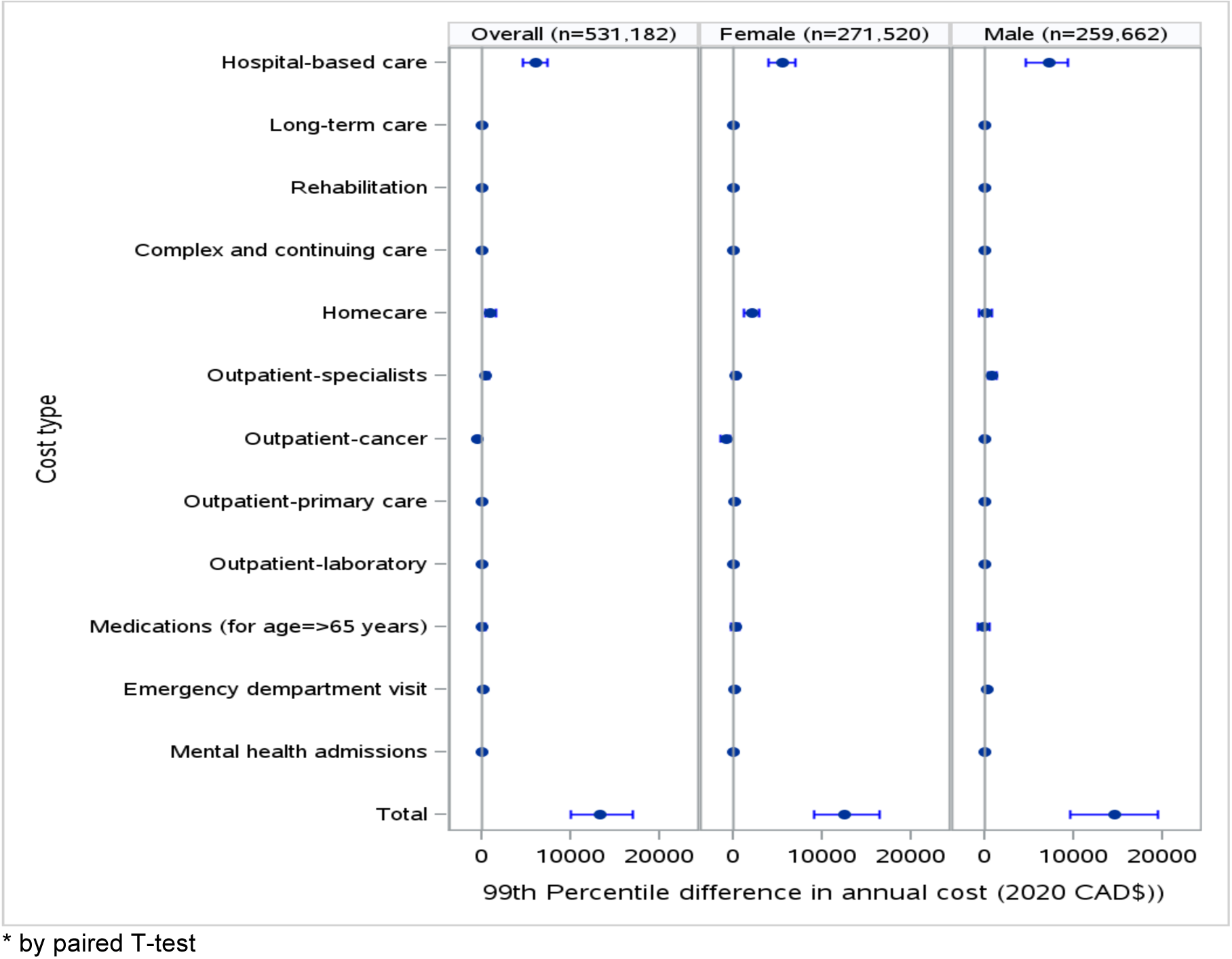
Differences in person-specific annual 99th percentile health care costs* for test-positive versus test-negative matched individuals, overall and by sex (95% confidence intervals^28^ in 2020 $CAD; starting 56 days after polymerase chain reaction test). Note range - $1,000 to +$20,000.

Mean person-specific total health care costs in the first year of follow-up were $487.08 (95% CI $393.69, $593.28) higher for test-positive compared to test-negative matched individuals. For test-positive females, mean person-specific total health care costs were $513.83 (95% CI $387.37, $638.40) higher in the first year of follow-up, and test-positive males had $459.10 (95% CI $304.60, $615.32) higher total health care costs compared to matched test-negative individuals.

At the 99^th^ percentile, test-positive individuals had $13,306.00 (95% CI, $10,004.00, $16,965.50) greater annual total health care costs compared to test-negative people, with an increase of $12,533.00 (95% CI % $9,008.50, $16,473.00) for test-positive females, and $14,604.00 (95% CI $9,565.50, $19,506.50) for test-positive males compared to their matched, test-negative counterparts.

### Component health care costs

The largest differences in mean component costs in the first year of follow-up for test-positive individuals were for hospital-based care ($244.83, 95% CI $188.59, $301.22), long-term care ($102.16, 95% CI $90.09, $113.89), and complex continuing care ($79.28, 95% CI $49.98, $107.96). There were also smaller but significantly higher costs for outpatient specialist, rehabilitation, emergency department, outpatient primary care (for females only, with no detected difference for males), and outpatient laboratory costs for test-positive individuals, and overall no detected differences in outpatient medication, mental health admission costs (no difference for males; decreased costs for test-positive females), or homecare (increased for females, no difference for males). Mean person-specific outpatient cancer care costs were lower (-$75.89, 95% CI -$96.71, -$54.45) over the first year of follow-up for test-positive individuals.

At the 99^th^ percentile, the largest cost differences between test-positive and test-negative individuals were for hospital care ($6035.00, 95% CI $4647.50, $7402.50) homecare ($937.00, 95% CI $389.50, $1,508.00; higher only for females), and outpatient specialist care ($407.00, 95% CI $177.00, $731.00; higher only for males). ED costs were $170.00 (95% CI $96.00, $241.00) higher for test-positive individuals, while there were no differences in costs for outpatient primary care, medications, or laboratory tests, and zero costs for mental health admission, complex continuing care, or long-term care. Outpatient cancer care costs were lower over the first year of follow-up by -$521 (95% CI -$708.50, -$399.00) overall, but test-positive females had -$844.00, (95% CI - $1,462.50, -$415.00) lower outpatient cancer care costs, with no detected difference for males.

### Censoring at long-term care entrance

A total of 1,157 people (827, 71.5% in the test-positive group) entered long-term care within six months of follow-up, and 1,752 (1,1165, 66.5% in the test-positive group) over 1-year follow-up. Results of sensitivity analyses censoring at entrance to long-term care were overall similar to the primary analyses, with the expected decreases in long-term care cost differences and resulting decrease in total costs (Supplemental Figure E4). Outpatient primary care costs and hospitalization cost differences were slightly lower than in the primary analyses.

## 6-month health care cost differences

Differences in annualized costs based on 6 month follow-up overall and stratified by sex are shown in Supplemental Figure E5. Magnitudes and patterns of person-specific total and component health care cost differences were similar for 6-month and 1-year follow-up, with the exceptions of long-term care, home care, outpatient medications, and outpatient primary care costs. Mean differences in person-specific long-term care and home care costs at 6-month follow-up were approximately half those for 1-year follow-up both males and females; for 6-month follow-up, test-positive males had higher mean outpatient primary care costs but no detected cost difference for 1-year follow-up. The magnitude of decrease in person-specific, mean outpatient cancer care costs were similar for 6-month and 1-year follow-up.

For 6-month follow-up, cost differences at the 99^th^ percentile generally similar in pattern and magnitude to those at 1-year follow-up, with the exceptions of outpatient primary, homecare, cancer care costs (for females) and outpatient medication costs (for males). At 6-months, test-positive females had higher outpatient primary care costs than test-negative females; at 1-year follow-up, there was no detected cost difference in outpatient primary care costs for females or males. Annualized 6-month homecare cost differences for females were nearly half those at 1-year follow-up, with a 6-month cost difference of $1,031.00 (95% CI $432.00, $1,504.00) and 1-year cost difference of $2,103.00 (95% CI $1,150.50, $2,939.50). For test-positive females, cancer care costs were lower than matched test-negative females, with a larger gap at 1-year follow up: at 6-month follow-up, the cost difference for cancer care was -$161.00 (95% CI -$353.00, $0.00); at 1-year follow-up, the cost-difference was -$844.00 (95% CI -$1,462.50, -$415.00). For males, 6-month outpatient medication costs were lower for test-positive versus test-negative males, with no detected difference at 1-year for males or females. For both 6-month and 1-year follow-up, 99^th^ percentile costs for long-term care, complex continuing care, and mental health hospitalizations were zero.

### Relative risks of post-acute high costs, any health care costs

RR for being a high-cost user of six component costs (outpatient cancer, rehabilitation, home care, mental health hospitalization, complex continuing care, and long-term care) were generally of smaller magnitude at 1-year follow-up than 6-month, with notable differences by sex (Table 2; Supplemental Table E5 for risk differences). For test-positive males, there was no detected difference in their RR of being a high-cost user of long-term care at either 6-month or 1-year follow-up; their 6-month RR were >1 for being high-cost users of outpatient care, rehabilitation, homecare, mental health hospitalization, and complex continuing care, but by 1-year follow-up only the RR of being a high-cost user of complex continuing care remained statistically significant. In contrast for test-positive females, 1-year RR for being high-cost users of outpatient cancer care, rehabilitation, mental health hospitalization, complex continuing care, and long-term care remained statistically significant, with smaller magnitude than 6-month RRs.

**Table 2:**
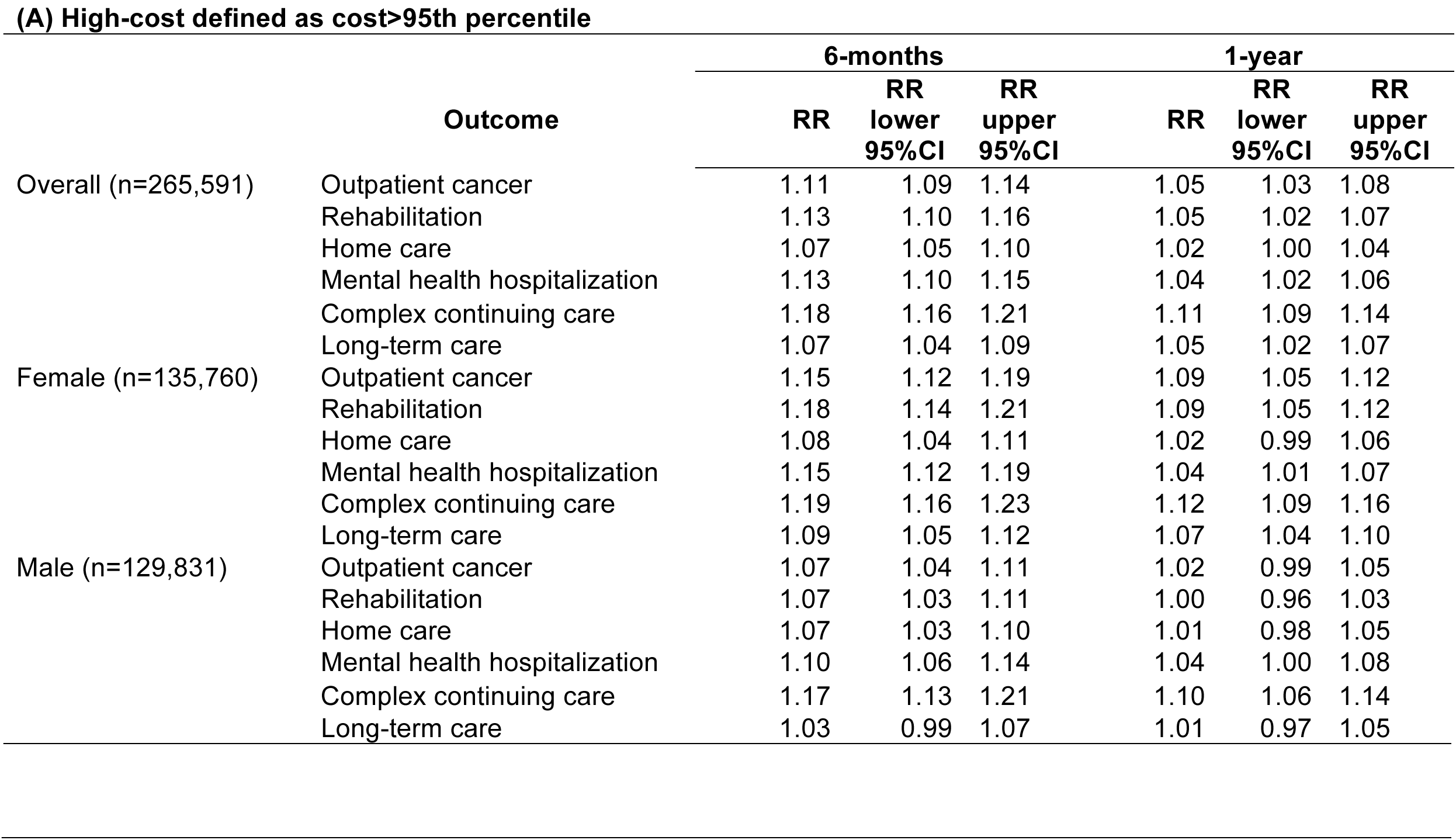
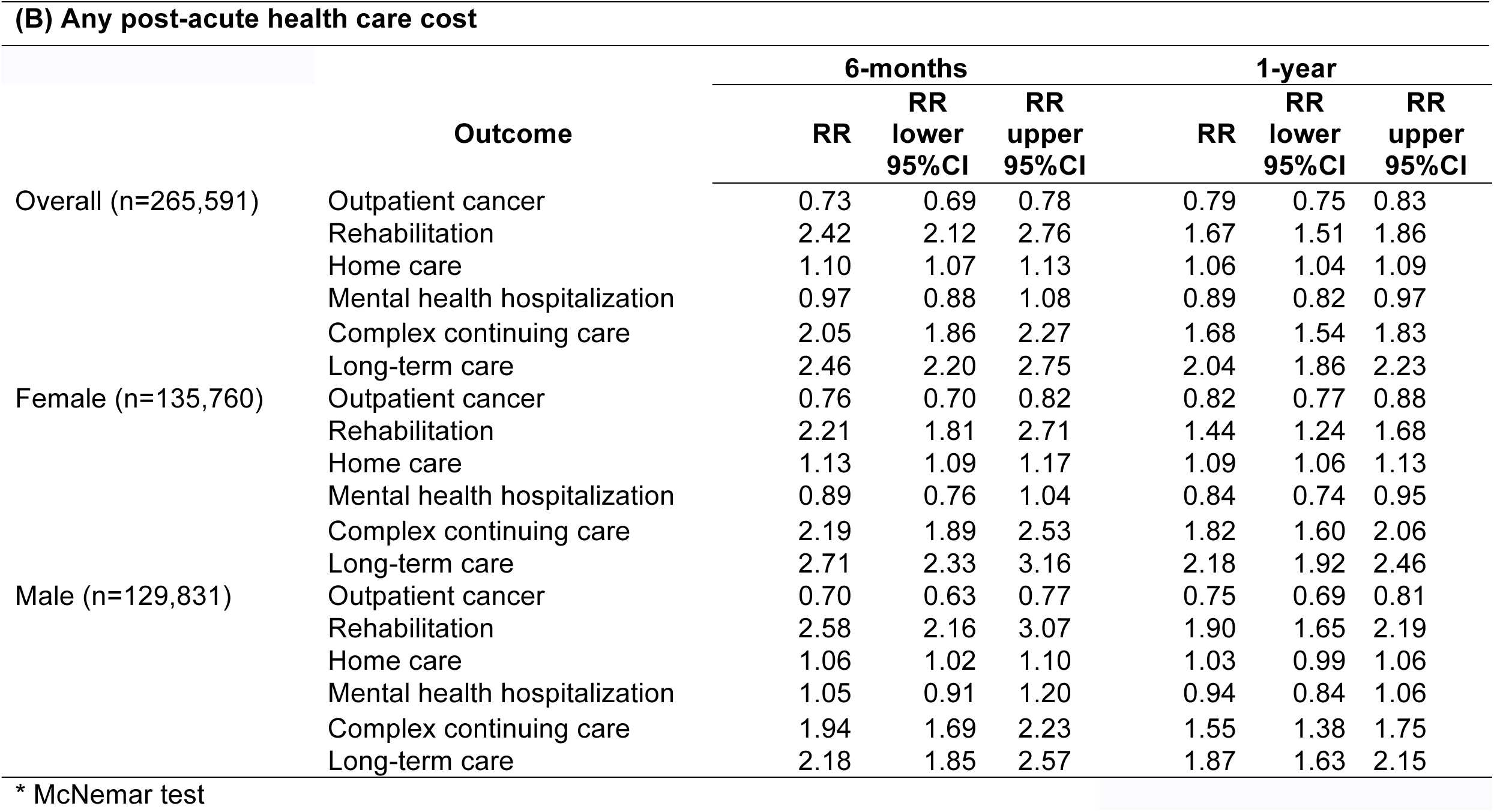
Relative risks* comparing test-positive versus test-negative people, overall and by sex, for: (A) being a highcost health care user (>95th percentile) within 6 months and 1 year of follow-up, and (B) any post-acute health care cost (>$0) within 6 months and 1 year of follow-up.

For any 6-month or 1-year post-acute cancer care, rehabilitation, home care, mental health hospitalization, complex continuing care, or long-term care, there was no detected difference for test-positive males compared to test-negative males. For test-positive females the risk was lower at 1-year compared to test-negative individuals. The RR for any outpatient cancer costs was lowest at 6-months and at 1-year follow-up remained lower compared to test-negative individuals for both males and females. RR of any spending on rehabilitation, home care, complex continuing care, and long-term care were highest for 6-month follow-up and generally decreased by 1-year but remained elevated compared to test-negative individuals. For males at 1-year there was no detected difference in the risk of any homecare costs compared to test-negative individuals.

## Discussion

Among 531,182 matched adults, post-acute (≥56 days after PCR testing) person-specific health care costs in the first year of follow-up were on average $487 higher ($513 for females, $459 for males) for individuals who had a positive SARS-CoV-2 PCR test compared to matched, test-negative individuals. Cost differences at the 99^th^ percentile over the first year of follow-up were $13,306 higher ($12,533 for females, $14,604 for males) for test-positive individuals. With annual mean health care expenditures of $4,800 per-person in Ontario for 2020,^29^ this represents >10% increase in mean health care costs over the first year of post-acute follow-up for test-positive individuals. In a conservative estimate based on a mean cost difference half that of our results and only half of adults in Ontario infected, this would translate to an estimated additional $1.52 billion health care costs over one year of post-acute follow-up. Such a conservative estimate does not account for costs that may extend beyond a year of post-acute follow-up or the potential for greater cost differences for re-infections.

The largest cost differences at the mean and 99^th^ percentile for both males and females was for hospital costs. At the mean, this was followed by long-term care and complex continuing care costs differences, while at the 99^th^ percentile the next largest cost differences were for homecare (for females), outpatient specialist (for males), and emergency department costs. We did not find evidence for increased costs for mental health hospitalizations, and for test-positive females mental health hospitalization costs were lower than matched test-negative women.

Primary care and emergency department mean costs were higher for test-positive individuals ($9.27 and $12.14, respectively). While person-specific dollar amounts for these cost differences are smaller compared to those for hospital ($243.00) or long-term care ($102.16), access to primary care is the lynchpin of the healthcare system,^30^ and >40% of adults seek emergency department care at least once a year.^31^ Thus, any increased primary care or emergency department costs following SARS-CoV-2 infection are likely to have clinically important consequences in a health care system already functioning beyond capacity, particularly if such an increase is sustained.^32^

Notably, outpatient mean cancer care costs were lower for test-positive individuals, and at the 99^th^ percentile outpatient cancer care costs were more than $800 lower for test-positive females over the first year of follow-up, with no detected difference for males. Overall, test-positive individuals were also less likely to have any outpatient cancer care costs, while test-positive women had increased risk of being a high-cost outpatient cancer care user over the first year of follow-up. Taken together, this raises the possibility that cancer diagnosis and/or treatment may have been impacted by SARS-CoV-2 infection, post-acute complications, or their impact on the healthcare system, and it is in line with previous work raising concerns about increases in undetected cancer cases since the start of the COVID-19 pandemic.^33^

We did not find that cost differences returned to baseline over the first year of follow-up. Similar total and component health care cost differences at 6-month and 1-year follow-up suggest sustained, increased health care costs following SARS-CoV-2 infection, even when follow-up began 56 days after a positive PCR.

Our findings are consistent with growing evidence that patient-reported post-acute symptoms and complications are common and persistent.^34–41^ An estimated 10% of people meet criteria for long COVID following SARS-CoV-2 infection, with incidence and prevalence varying by case definition, outcome measures, pandemic phase, vaccination status, and follow-up duration.^42–44^ Two studies have addressed post-acute COVID-19 health care costs, though both were limited to 6-month follow-up of populations covered by specific health insurance programs early in the pandemic, used diagnosis codes to identify COVID-19 cases, and they were conducted in the U.S., which has the highest health care costs in the world.^45–47^ Six-month health care costs (i.e., money paid by insurance companies for provision of medical care) for commercially insured patients <65 years of age were 1.46 times higher than matched controls,^45^ and compared to pre-pandemic controls, mean cost differences for people after COVID-19 were higher and driven largely by hospital costs: mean cost difference $763/month USD for commercially insured patients; mean cost difference $2,337/month USD for Medicare patients.^46^ One additional study in the US examined condition-specific health care utilization in the year following a clinical diagnosis of COVID-19, and also found increased risk of mortality and health care utilization for a range of disease conditions.^47^ These likely underestimate total health care use and cost differences, as home-based and long-term medical care is often not covered by health insurance in the U.S.

Our population-level data for a large, universal health care system reduces risks of selection bias and differential follow-up. Use of health care costs rather than diagnostic codes or patient-reported symptoms reduces the risk of recall bias, and our findings are likely generalizable to other similar universal health care systems. We examined the totality of publically funded health care costs, which includes home health aids, home visits by nurses, assisted living, rehabilitation, and nursing home care as well as hospital, clinic, and outpatient medications costs, and we were able to examine outpatient costs for primary care, specialist, and cancer care separately. Use of PCR to identify cases likely biases our results towards the null by underestimating true burden infection,^48, 49^ but we chose a highly specific case definition in order to fully examine post-acute health care costs with less risk of erroneously classifying people as having had SARS-CoV-2 infection. Our findings may further have been biased towards the null by likely overrepresentation of ill people in the test-negative population, e.g., patients undergoing repeated PCR testing for chemotherapy, dialysis, etc. Our findings indicate that in addition to higher mean post-acute health care costs, a small proportion of individuals had markedly higher health care costs following infection, and while hospital costs were the main driver of total cost, important costs were also incurred in settings other than the hospital or outpatient clinics.

We also add to existing literature regarding sex differences in COVID-19^50–57^ by identifying sex differences for component health care costs. Test-positive males had higher mean cost differences for rehabilitation and mental health care, while test-positive women had higher mean cost differences for outpatient medications, primary care, homecare, and long-term care. Further work is needed to examine the interplay between social factors (e.g., caregiver availability for men versus women) and biological factors in order to appropriate plan for and allocate long COVID resources.

With few protective measures in place to prevent ongoing airborne spread and many people therefore expected to contract one or more SARS-CoV-2 infection annually,^38^ sustained health care cost increases of the magnitude we describe will require significant health care system restructuring, innovation, and investment of resources, particularly for health systems with existing prolonged wait times to access care, insufficient supply of acute and long-term care beds, or projected loss of health care workers.^58–66^ Health care workers appear to have higher risk of infection and developing post-acute complications, with 18% unable to return to pre-infection clinical workloads, including nearly half of whom were infected after been fully vaccinated.^67–69^ Further, our findings do not account for costs related to disability, unemployment, or reduced quality of life after acute COVID-19, which has been reported to be comparable to that of stage 4 lung cancer.^67, 70–74^ Implementing protections and other measures to prepare for post-acute complications of SARS-CoV-2 infection could forestall considerably avoidable health care and other costs.^75^

### Limitations

We compared individuals with positive PCR tests to those with negative test results and matched on myriad baseline healthcare access and use, thus our findings may not generalize to people who do not have similar access to medical care. To address time trends in testing and changes to the health care system during early phases of the pandemic,^58, 76–78^ we used both propensity score matching and hard matching by sex/pregnancy status, 2-week time blocks, public health unit, and vaccination status. Despite extensive matching including multiple socio-demographic and clinical factors, there is potential residual confounding by unmeasured variables such as body mass index or symptoms severity and duration, although control individuals may also not have had symptoms. However, growing evidence suggests that even so-called mild infections (e.g., those caused by omicron variants or that did not result in hospitalization) are associated with important post-acute complications that, combined with the large number of infections, have important health system implications.^79^ Questions also remain whether healthcare costs remain elevated beyond a year of follow-up, as well as whether vaccine recency and reinfection may modify the trajectory and duration of post-acute complications.^80–84^ This study also cannot address health care needs that did not result in seeking medical care or health care costs incurred outside of the traditional health care system.

## Conclusions

Over the first year of post-acute follow-up, health care costs after a positive SARS-CoV-2 PCR test are significantly higher compared to matched test-negative individuals, with the largest increases in mean person-specific health care costs for hospitalization, long-term care and complex continuing care, while increases in the 99^th^ percentile of costs were driven by hospitalizations, outpatient specialists (for males), and homecare costs (for women). Outpatient cancer care costs were lower, raising concerns about potential delays in diagnosis or treatment. Differences in health cares costs were similar at 6-month and 1-year follow-up. These findings can be used by policymakers to anticipate and plan for post-acute health care costs following SARS-CoV-2 infection.

## Data Availability

All data produced in the present study are available upon reasonable request to the authors and within local legal regulations for data security and management. Please contact ICES for details: https://www.ices.on.ca

## Acknowledgments

This study was supported by ICES, which is funded in part by an annual grant from the Ontario Ministry of Health and the Ministry of Long-Term Care (MOHLTC). Part of this material is based on data and/or information compiled and provided by the Canadian Institute for Health Information (CIHI). The authors acknowledge that the clinical registry data used in this publications is from participating hospitals through CorHealth Ontario, which serves as an advisory body to the MOHLTC, is funded by the MOHLTC, and is dedicated to improving the quality, efficiency, access and equity in the delivery of the continuum of adult cardiac, vascular and stroke services in Ontario, Canada. The authors thank IQVIA Solutions Canada Inc. for use of their Drug Information File. Parts of this report are based on Ontario Registrar General (ORG) information on deaths, the original source of which is ServiceOntario. This study was supported by the Ontario Health Data Platform (OHDP), and Province of Ontario initiative to support Ontario’s ongoing response to COVID-19 and its related impacts. Parts of this material are based on data and/or information compiled and provided by CIHI and Cancer Care Ontario (CCO). The analyses, results, conclusions, opinions, and statements reported are those of the authors and are independent of the data and funding sources. No endorsements by ICES, the Ontario MOHLTC, CIHI, OHDP or its partners, ORG or the Ministry of Government Services, CCO, or the Province of Ontario is intended or should be inferred.

Dr. McNaughton is supported by the Sunnybrook Research Institute, the Practice Plan of the Department of Emergency Services at Sunnybrook Health Sciences Centre, and the University of Toronto.

Dr. Atzema is supported by the Sunnybrook Research Institute, the Practice Plan of the Department of Emergency Services at Sunnybrook Health Sciences Centre, and by a Mid-Career Investigator Award from the Heart and Stroke Foundation.

