## Supplement for "Post-acute health care costs following SARS-CoV-2 infection: A retrospective cohort study of among 531,182 matched adults"

### Supplemental Materials

**Supplemental Table E1.** Databases used for cohort construction and variable definitions

**Supplemental Table E2.** Variables used propensity score matching and hard matching

**Supplemental Table E3:** Interaction by sex (propensity score-matched cohort)

**Supplemental Figure E1.** Cohort construction

**Supplemental Table E4**. Cohort characteristics before propensity score matching

**Supplemental Figure E2:** Difference in annual median health care costs (95% confidence intervals in 2020 $CAD, starting 56 days after polymerase chain reaction test) for test-positive versus test-negative matched individuals, overall and by sex

**Supplemental Figure E3:** Difference in annual 95th percentile health care costs (95% confidence intervals in 2020 $CAD, starting 56 days after polymerase chain reaction test) for test-positive versus test-negative matched individuals, overall and by sex

**Supplemental Figure E4**: Sensitivity analyses censored at entrance to long-term care

**Supplemental Figure E5:** Differences in 6-month health care costs (95% confidence intervals in 2020 $CAD, starting 56 days after polymerase chain reaction test) for test-positive versus test-negative matched individuals, overall and by sex, at the following distributions: (A) mean, (B) median, (C) 95$^{th}$ percentile, (D) 99$^{th}$ percentiles

**Supplemental Table E5:** Risk differences between test-positive and test-negative people for being a high-cost user within 6 months and 1 year, overall and by sex, defined as (A) >95th percentile and (B) any post-acute health care cost ($>0)

**Supplemental Table E1.** Databases used for cohort construction and variable definitions. Additional details are available at https://datadictionary.ices.on.ca/Applications/DataDictionary/Default.aspx

| Database | Cost Components | Costing Methods (standardized to 2020CAD) |
|---|---|---|
| Ontario Health Insurance Plan (OHIP) | Outpatient physician visits<br>Laboratory services<br>Non-physician services | -Fee-for-service + 'shadow billing'<br>-Technical + professional fees |
| National Ambulatory Care Reporting System (NACRS) | Emergency department visits<br>Dialysis visits<br>Oncology clinic visits | Resource weight intensity x cost/weighted case |
| Discharge Abstract Database (DAD) | Inpatient hospitalizations | Resource weight intensity x cost/weighted case; as appropriate, prorated to exclude days prior to start of follow up |
| Client Agency Program Enrolment | Capitation costs | Base rate x age-sex multiplier + applicable premiums |
| Ontario Drug Benefit (ODB) | Medication use | Total payments |
| National Rehabilitation Reporting System | Rehabilitation admissions | Rehabilitation patient group x rehabilitation cost-weight |
| Continuing Care Reporting System | Complex continuing care admissions<br>Long-term care | -Case mix index x cost/weighted day<br>-Length of stay x cost/day |
| Home Care Database | Home care services | Standard cost per service |
| Mental Health Reporting System | Mental health admissions | (Length of stay x case mix index) x cost /weighted day |
| Same Day Surgery | Same day surgery | Resource weight intensity x cost/weighted case |

| **Other Databases** | |
|---|---|
| Registered Persons Database (RPDB) | Congestive Heart Failure (CHF) |
| Canadian Institute for Health Information (CIHI) | Ontario Asthma dataset (ASTHMA) |
| Yearly Health Services Contact | Chronic Obstructive Pulmonary Disease (COPD) |
| Ontario Marginalization Index (ONMARG) | Ontario Cancer Registry |
| Ontario COVID-19 Vaccine Data | Ontario Dementia Database (DEMENTIA |
| COVID19 Integrated Testing Data | ICES Physician Database |
| Ontario Laboratories Information System | Corporate Provider Database |
| Ontario Hypertension Dataset (HYPER) | Ontario Diabetes Dataset (DIABETES) |

| **Descriptions of specialized health care services** | |
|---|---|
| Homecare | Providing of services for people of all ages who require care in their home, school, or in the community, and seniors with complex medical conditions who otherwise would be able to live in their home. Includes health professional home visits (e.g., nursing care, physio/physical therapy, occupational therapy, speech-language therapy, social work, nutrition support, home healthcare supplies, personal care support for activities of daily living, homemaking, and family-managed home care/self-directed care, and end-of-life care at home. |
| Complex continuing care | Complex Continuing Care (CCC) provides continuing, medically complex and specialized services to people of all ages, sometimes over extended periods of time. CCC is offered in healthcare centres for people who have long-term illnesses or disabilities typically requiring skilled, technology-based care not available at home or in long-term care facilities. Patients must be medically stable and are often eligible if they have an uncuffed tracheostomy, peripherally inserted central catheter, feeding tube, or nephrostomy tube. |
| Rehabilitation | Hospital beds for medically complex patients provide |

| | |
|---|---|
| | rehabilitation to gradually improve strength and movement. Care includes access to doctors or nurse practitioners, nursing, and physio (physical) and occupational therapy. Typical short-term stays are up to three months. Long-term patients are understood to be permanent residents unless conditions change. |
| Long-term care | Similar to nursing home care for medically complex patients in the United States. Long-term care homes provide homecare and services for people whose needs cannot be met in the community. Monthly out-of-pocket patient costs are determined by the Ministry and as of July 1, 2023 range from $1,986.92-$2,838.49 per month. |

**Supplemental Table E2.** Variables used propensity score matching and hard matching. Hard matched variables indicated by*: sex, outbreak period, and 1-year baseline total healthcare costs. Please see Supplemental Table E1 for datasets.

| Variable | Definition and/or source |
|---|---|
| Age | Years, restricted cubic spline |
| Sex* | Male, female (yes/no pregnant) |
| Outbreak period* | Two week blocks, beginning January 28, 2020 |
| Public health unit | 34 units |
| Neighbourhood income | Quintiles |
| Residential instability[1] | Quintiles, Ontario Marginalization Index; e.g., percentage living alone, dwellings not owned |
| Material deprivation[1] | Quintiles, Ontario Marginalization Index; e.g., percentage unemployed, without a high school degree |
| Dependency[1] | Quintiles, Ontario Marginalization Index; e.g., percentage of seniors, individuals not participating in the labour force |
| Ethnic concentration[1] | Quintiles, Ontario Marginalization Index; e.g., percentage of recent immigrants, those who self-identify as a "visible minority |
| Rurality | Population <10,000 |
| Diabetes | ICES DIABETES |
| Pregnancy | ICES MOMBABY[2-5] |
| Hypertension | ICES HYPER |
| Acute myocardial infarction | ≥1 DAD ICD10: I21, I22, I25.2 |
| Percutaneous coronary intervention | CIHI DAD/SDS: CCP: 48.02, 48.09; CCI: 1IJ50, 1IJ57GQ 1IJ80, 1IJ26, 1IJ54, 1IJ55; OHIP: Z434, Z448, Z449, Z460, Z461 |
| Coronary artery bypass surgery | (CIHI DAD/SDS): CCI 1IJ76; CCP 48.1, 48.2; OHIP R742, R743 |
| Ischemic stroke | One DAD, 2 OHIP, or 1 NACRS&1OHIP: ICD-10 codes I63, I64, H341 (excluding I63.6) as ANY diagnosis type, exclude suspect; ICD-9 code 434, 436 for OHIP |
| Haemorrhagic stroke | 1 record in DAD or NACRS: ICD-10 I60, I61; ICD-9 430, 431 |
| Major bleeding event | ICD 10:<br>GI: I850, I983, K250/252/254/256, K260/262/264/266, K270/272/274/276, K280/282/284/286, K290, K661, K920, K921, K922<br>ICH: I60, I61, I620, I621, I629<br>GU: N020-029, R310,R311, R318<br>Resp: R040, R041, R042, R048, R049<br>Other: R58, D68.3, H35.6, H45.0, M25.0 |
| Solid cancer, hematologic cancer | OCR: OCR_TOPOG_CD, OCR_DIAG_DATE |
| Same-day surgery in prior 6 weeks | SDS_ADMDATE, SDS_INCODE1-10, SDS_CACSANETECH |
| Valvular disease | ICD9: 394, 395, 396<br>ICD10: I019, I020, I05, I08, I099, I342, I348, I349<br>ICD10 code Z952, and CCI codes 1HS90LACF, 1HT90LACF, 1HU90DACF, 1HU90LACF, 1HU90PNCF, 1HV90LACF, 1HV90LACFA, 1HV90LACFL, 1HV90LACFN, 1HV90WJCFN |
| Emphysema | COPD_SPECIFIC in COPD database |
| Asthma | ASTHMA_SPEC in ASTHMA database |
| Atrial fibrillation | Any of the following:<br>-history of hospitalization (CIHI DAD)[6]: ICD9 427.3 or ICD10 I48 as any diagnosis type, including suspected<br>-history of ED visit with same codes<br>-4 OHIP claims in 1 year (OHIP) *dxcode* 427 |
| Heart failure | CHF database |
| Ischemic heart disease | PCI, CABG, (1 HOSP in DAD with any codes I20-I25) or (2 OHIP billings within a one-year period with dx codes 410-414) |
| Renal disease | DAD code: ICD10 codes E102, E112, E132, E142, I12, I13, N01.*, N03.*, N05.*, N08.*, N18.*, N19.*, N25.*<br>or<br>Chronic Dialysis (Any 2 codes within 90 days of one another):<br>OHIP: R849, R850, G323, G325, G326, G330, G331, G860, G333, G083, G091, G085, G295, G082, G090, G092, G093, G094, G861, G862, G863, G864, G865, G866, G294, G095, G096 CCP: 51.95, 66.98 |

| | |
|---|---|
| | NACRS: CCI: 1PZ21HQBR, 1PZ21HPD4 |
| Pneumonia | (CIHI DAD, NACRS, OHIP) – ICD-10 codes J10.0, J11.0 or J12-J18 as ANY diagnosis type, exclude suspected[7]; OHIP dxcode 486, excluding claims associated with fee codes G538, G539, G840-G848, G590, G591 or G700 (administration of vaccinations) |
| Dementia | (1 DAD or 3 OHIP billings separated by 30 days, within a 2-year period or any cholinesterase inhibitor from ODB)<br>ICD10 codes F00-F03, F051, G30, G31, R54<br>ICD9 code 290, 294, 331, 797<br>SUBCLNAM= CHOLINESTERASE INHIBITORS<br>ODB: donepezil, galantamine, or rivastigmine (DIN: 02232043, 02232044, 02269457, 02269465, 02244298, 02244299, 02244300, 02244302, 02266717, 02266725) or Tacrine (Cognex) (DIN: 66123288, 66123290, 66123306, 66123318) |
| Alcohol substance use disorder | ICD10 codes F1094, F1029, F1019, F1099, F10250, F10150, F10950, F10920, F10929, F10251, F10251, F10151, F10951, F1027, F1097, F1026, F1096, F10129, F10120, F1010, F1021, F1020, F10220, F10229<br>ICD9 codes 291, 303 |
| Aggregated Diagnosis Group category | Aggregated Diagnosis Group (ADG) category group (from OHIP, NACRS-ED, CIHI DAD). The Johns Hopkins ACG© System Version 10.0 was used. |
| John Hopkins frailty indicator | (OHIP, NACRS-ED CIHI DAD) |
| Influenza vaccination | 2019-20 influenza vaccination (OHIP, ODB) – Received between Sep 1, 2019 and COVID test date. Note: does not capture vaccinations received outside doctor's offices and pharmacies |
| Venous thromboembolism | CIHI-NACRS, DAD, OHIP<br>ICD-10 I26.*, I26.0, I26.9, I80.* (excluding I80.0), I80.1, I80.2 (excluding I82.0, I82.1, I82.3), I80.3, I80.8, I80.9, I82.*, I82.2, I82.8, I82.9,<br>OHIP DXCODE 415, 451, 452, 453 |
| Mental health ED visit | OHIP[8-10] |
| MHA Outpatient Services | OHIP[8-10] |
| 1-year baseline total healthcare costs* | See Supplemental Table E1 |
| * variable used for hard matching | |

**Supplemental Material References**

**Supplemental Table E3**. Interaction by sex (tested in matched cohort)

| Component cost | P-value |
|---|---|
| Total | 0.5908 |
| Hospital | 0.4838 |
| Emergency department | 0.5389 |
| Outpatient medication | **0.0739*** |
| Outpatient specialists | 0.5944 |
| Outpatient cancer | 0.2219 |
| Outpatient primary care | **0.003*** |
| Outpatient laboratory | **<.0001*** |
| Rehabilitation care | **0.0008*** |
| Homecare | **0.0037*** |
| Mental health | **0.0225*** |
| Complex continuing care | 0.8936 |
| Long-term care | **<.0001*** |

* Significant, at P-value threshold of 0.10

Supplemental Figure E1: Cohort construction

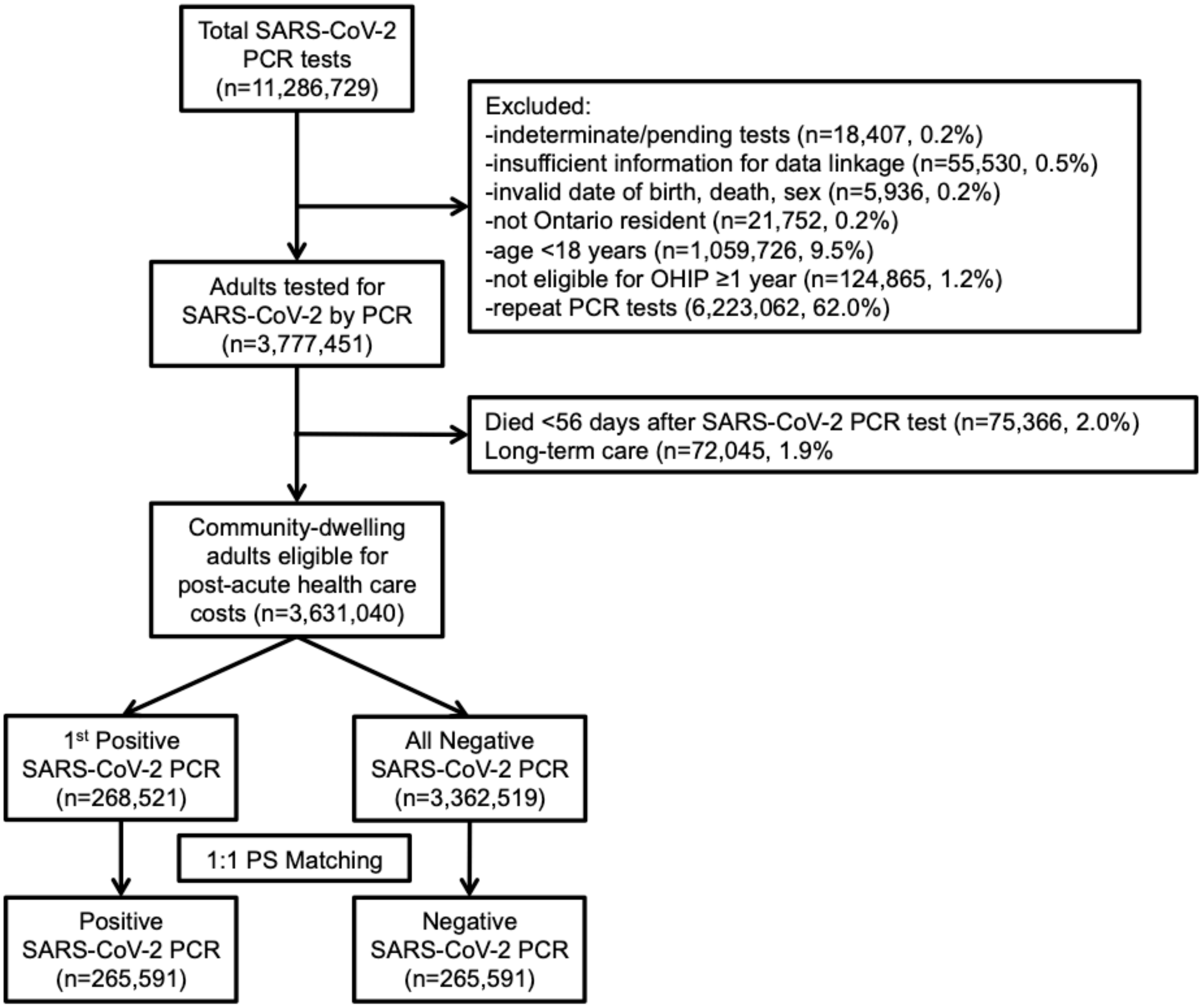

Abbreviations: PCR, polymerase chain reaction; OHIP, Ontario Health Insurance Plan; PS, propensity score

**Supplemental Table E4.** Cohort characteristics before propensity score matching

| | | Total | Test Negative | Test Positive | Standardized Difference |
|---|---|---|---|---|---|
| | | N=3,631,040 | N=3,362,519 | N=268,521 | |
| Age (years) | Mean (SD) | 46.58 (18.11) | 46.77 (18.17) | 44.18 (17.18) | 0.146 |
| | Median (Q1-Q3) | 45 (31-60) | 45 (31-60) | 43 (29-56) | 0.141 |
| Sex | F - n (%) | 1,970,075 (54.3%) | 1,834,685 (54.6%) | 135,390 (50.4%) | 0.083 |
| | M - n (%) | 1,629,552 (44.9%) | 1,498,276 (44.6%) | 131,276 (48.9%) | 0.087 |
| | P - n (%) | 31,413 (0.9%) | 29,558 (0.9%) | 1,855 (0.7%) | 0.021 |
| Rural | Missing Data - n (%) | 9,451 (0.3%) | 8,780 (0.3%) | 671 (0.2%) | 0.002 |
| | Y - n (%) | 350,523 (9.7%) | 339,985 (10.1%) | 10,538 (3.9%) | 0.244 |
| Johns Hopkins Frailty Indicator | n (%) | 123,999 (3.4%) | 116,104 (3.5%) | 7,895 (2.9%) | 0.029 |
| Adjusted Clinical Group Categories | | | | | |
| Aggregated Diagnosis Group Category 1 | n (%) | 2,782,083 (76.6%) | 2,574,005 (76.5%) | 208,078 (77.5%) | 0.022 |
| Aggregated Diagnosis Group Category 2 | n (%) | 2,731,200 (75.2%) | 2,529,253 (75.2%) | 201,947 (75.2%) | 0 |
| Aggregated Diagnosis Group Category 3 | n (%) | 2,235,629 (61.6%) | 2,073,361 (61.7%) | 162,268 (60.4%) | 0.025 |
| Aggregated Diagnosis Group Category 4 | n (%) | 195,459 (5.4%) | 182,201 (5.4%) | 13,258 (4.9%) | 0.022 |
| Aggregated Diagnosis Group Category 5 | n (%) | 1,085,536 (29.9%) | 1,020,361 (30.3%) | 65,175 (24.3%) | 0.137 |
| Aggregated Diagnosis Group Category 6 | n (%) | 1,523,716 (42.0%) | 1,407,922 (41.9%) | 115,794 (43.1%) | 0.025 |
| Aggregated Diagnosis Group Category 7 | n (%) | 193,342 (5.3%) | 181,715 (5.4%) | 11,627 (4.3%) | 0.05 |
| Aggregated Diagnosis Group Category 8 | n (%) | 240,396 (6.6%) | 224,682 (6.7%) | 15,714 (5.9%) | 0.034 |
| Aggregated Diagnosis Group Category 9 | n (%) | 339,984 (9.4%) | 316,552 (9.4%) | 23,432 (8.7%) | 0.024 |
| Aggregated Diagnosis Group Category 10 | n (%) | 1,263,741 (34.8%) | 1,185,427 (35.3%) | 78,314 (29.2%) | 0.131 |
| Aggregated Diagnosis Group Category 11 | n (%) | 1,183,663 (32.6%) | 1,103,389 (32.8%) | 80,274 (29.9%) | 0.063 |
| Aggregated Diagnosis Group Category 12 | n (%) | 165,525 (4.6%) | 152,992 (4.5%) | 12,533 (4.7%) | 0.006 |
| Recent flu vaccination | n (%) | 1,130,869 (31.1%) | 1,064,518 (31.7%) | 66,351 (24.7%) | 0.155 |
| Hypertension | n (%) | 877,852 (24.2%) | 816,144 (24.3%) | 61,708 (23.0%) | 0.03 |
| Diabetes | n (%) | 433,350 (11.9%) | 395,575 (11.8%) | 37,775 (14.1%) | 0.069 |
| Emphysema | n (%) | 97,824 (2.7%) | 93,686 (2.8%) | 4,138 (1.5%) | 0.086 |
| Heart failure | n (%) | 93,663 (2.6%) | 88,669 (2.6%) | 4,994 (1.9%) | 0.052 |
| Dementia | n (%) | 45,684 (1.3%) | 42,410 (1.3%) | 3,274 (1.2%) | 0.004 |
| Asthma | n (%) | 442,882 (12.2%) | 414,612 (12.3%) | 28,270 (10.5%) | 0.057 |
| Cancer | n (%) | 116,997 (3.2%) | 112,188 (3.3%) | 4,809 (1.8%) | 0.098 |
| Surgery in previous 6 weeks | n (%) | 59,924 (1.7%) | 57,709 (1.7%) | 2,215 (0.8%) | 0.08 |
| Ischemic stroke | n (%) | 45,455 (1.3%) | 42,908 (1.3%) | 2,547 (0.9%) | 0.031 |
| Hemorrhagic stroke | n (%) | 3,757 (0.1%) | 3,533 (0.1%) | 224 (0.1%) | 0.007 |
| Valvular disease | n (%) | 4,844 (0.1%) | 4,609 (0.1%) | 235 (0.1%) | 0.015 |

| | | | | | |
|---|---|---|---|---|---|
| Ischemic heart disease | n (%) | 177,360 (4.9%) | 167,514 (5.0%) | 9,846 (3.7%) | 0.065 |
| Major bleeding | n (%) | 39,366 (1.1%) | 37,210 (1.1%) | 2,156 (0.8%) | 0.031 |
| Renal disease | n (%) | 46,728 (1.3%) | 43,949 (1.3%) | 2,779 (1.0%) | 0.025 |
| Pneumonia | n (%) | 291,202 (8.0%) | 271,036 (8.1%) | 20,166 (7.5%) | 0.021 |
| Alcohol use disorder | n (%) | 29,427 (0.8%) | 27,759 (0.8%) | 1,668 (0.6%) | 0.024 |
| Venous thromboembolism | n (%) | 420,456 (11.6%) | 396,371 (11.8%) | 24,085 (9.0%) | 0.093 |
| | Missing Data - n (%) | 10,824 (0.3%) | 10,050 (0.3%) | 774 (0.3%) | 0.002 |
| Income Quintile 1 | n (%) | 697,116 (19.2%) | 630,532 (18.8%) | 66,584 (24.8%) | 0.147 |
| Income Quintile 2 | n (%) | 707,319 (19.5%) | 649,204 (19.3%) | 58,115 (21.6%) | 0.058 |
| Income Quintile 3 | n (%) | 729,395 (20.1%) | 672,162 (20.0%) | 57,233 (21.3%) | 0.033 |
| Income Quintile 4 | n (%) | 731,678 (20.2%) | 684,228 (20.3%) | 47,450 (17.7%) | 0.068 |
| Income Quintile 5 | n (%) | 754,708 (20.8%) | 716,343 (21.3%) | 38,365 (14.3%) | 0.184 |
| | Missing Data - n (%) | 39,428 (1.1%) | 37,365 (1.1%) | 2,063 (0.8%) | 0.036 |
| Instability Quintile 1 (Least unstable) | n (%) | 763,550 (21.0%) | 691,223 (20.6%) | 72,327 (26.9%) | 0.15 |
| Instability Quintile 2 | n (%) | 671,203 (18.5%) | 626,597 (18.6%) | 44,606 (16.6%) | 0.053 |
| Instability Quintile 3 | n (%) | 650,977 (17.9%) | 610,179 (18.1%) | 40,798 (15.2%) | 0.079 |
| Instability Quintile 4 | n (%) | 654,660 (18.0%) | 610,967 (18.2%) | 43,693 (16.3%) | 0.05 |
| Instability Quintile 5 (Most unstable) | n (%) | 851,222 (23.4%) | 786,188 (23.4%) | 65,034 (24.2%) | 0.02 |
| | Missing Data - n (%) | 39,428 (1.1%) | 37,365 (1.1%) | 2,063 (0.8%) | 0.036 |
| Deprivation Quintile 1 (Least deprived) | n (%) | 848,007 (23.4%) | 802,901 (23.9%) | 45,106 (16.8%) | 0.177 |
| Deprivation Quintile 2 | n (%) | 755,915 (20.8%) | 708,278 (21.1%) | 47,637 (17.7%) | 0.084 |
| Deprivation Quintile 3 | n (%) | 684,713 (18.9%) | 632,209 (18.8%) | 52,504 (19.6%) | 0.019 |
| Deprivation Quintile 4 | n (%) | 650,256 (17.9%) | 594,666 (17.7%) | 55,590 (20.7%) | 0.077 |
| Deprivation Quintile 5 (Most deprived) | n (%) | 652,721 (18.0%) | 587,100 (17.5%) | 65,621 (24.4%) | 0.172 |
| | Missing Data - n (%) | 39,428 (1.1%) | 37,365 (1.1%) | 2,063 (0.8%) | 0.036 |
| Dependency Quintile 1 (Least dependent) | n (%) | 993,323 (27.4%) | 901,394 (26.8%) | 91,929 (34.2%) | 0.162 |
| Dependency Quintile 2 | n (%) | 731,303 (20.1%) | 671,373 (20.0%) | 59,930 (22.3%) | 0.058 |
| Dependency Quintile 3 | n (%) | 626,297 (17.2%) | 582,059 (17.3%) | 44,238 (16.5%) | 0.022 |
| Dependency Quintile 4 | n (%) | 594,603 (16.4%) | 557,097 (16.6%) | 37,506 (14.0%) | 0.072 |
| Dependency Quintile 5 (Most dependent) | n (%) | 646,086 (17.8%) | 613,231 (18.2%) | 32,855 (12.2%) | 0.168 |
| | Missing Data - n (%) | 39,428 (1.1%) | 37,365 (1.1%) | 2,063 (0.8%) | 0.036 |
| Ethnic Concentration Quintile 1 (Least) | n (%) | 552,466 (15.2%) | 535,172 (15.9%) | 17,294 (6.4%) | 0.304 |
| Ethnic Concentration Quintile 2 | n (%) | 604,881 (16.7%) | 579,980 (17.2%) | 24,901 (9.3%) | 0.237 |
| Ethnic Concentration Quintile 3 | n (%) | 670,672 (18.5%) | 635,538 (18.9%) | 35,134 (13.1%) | 0.159 |
| Ethnic Concentration Quintile 4 | n (%) | 783,968 (21.6%) | 726,900 (21.6%) | 57,068 (21.3%) | 0.009 |
| Ethnic Concentration Quintile 5 (Most) | n (%) | 979,625 (27.0%) | 847,564 (25.2%) | 132,061 (49.2%) | 0.512 |
| Public health unit | Missing Data - n (%) | 9,838 (0.3%) | 9,146 (0.3%) | 692 (0.3%) | 0.003 |
| | 2226 - n (%) | 26,512 (0.7%) | 26,287 (0.8%) | 225 (0.1%) | 0.106 |
| | 2227 - n (%) | 38,987 (1.1%) | 37,168 (1.1%) | 1,819 (0.7%) | 0.046 |
| | 2230 - n (%) | 162,703 (4.5%) | 151,132 (4.5%) | 11,571 (4.3%) | 0.009 |
| | 2233 - n (%) | 36,617 (1.0%) | 35,864 (1.1%) | 753 (0.3%) | 0.096 |

| | | | | | |
|---|---|---|---|---|---|
| | 2234 - n (%) | 24,956 (0.7%) | 23,709 (0.7%) | 1,247 (0.5%) | 0.032 |
| | 2235 - n (%) | 50,556 (1.4%) | 49,540 (1.5%) | 1,016 (0.4%) | 0.115 |
| | 2236 - n (%) | 145,938 (4.0%) | 137,409 (4.1%) | 8,529 (3.2%) | 0.049 |
| | 2237 - n (%) | 146,639 (4.0%) | 136,426 (4.1%) | 10,213 (3.8%) | 0.013 |
| | 2238 - n (%) | 43,705 (1.2%) | 43,262 (1.3%) | 443 (0.2%) | 0.132 |
| | 2239 - n (%) | 12,439 (0.3%) | 12,084 (0.4%) | 355 (0.1%) | 0.046 |
| | 2240 - n (%) | 30,100 (0.8%) | 28,752 (0.9%) | 1,348 (0.5%) | 0.043 |
| | 224 n (%) | 55,293 (1.5%) | 54,644 (1.6%) | 649 (0.2%) | 0.144 |
| | 2242 - n (%) | 37,243 (1.0%) | 34,901 (1.0%) | 2,342 (0.9%) | 0.017 |
| | 2243 - n (%) | 45,285 (1.2%) | 44,218 (1.3%) | 1,067 (0.4%) | 0.1 |
| | 2244 - n (%) | 119,934 (3.3%) | 114,752 (3.4%) | 5,182 (1.9%) | 0.092 |
| | 2246 - n (%) | 115,348 (3.2%) | 108,164 (3.2%) | 7,184 (2.7%) | 0.032 |
| | 2247 - n (%) | 31,184 (0.9%) | 30,843 (0.9%) | 341 (0.1%) | 0.11 |
| | 2249 - n (%) | 24,286 (0.7%) | 23,711 (0.7%) | 575 (0.2%) | 0.073 |
| | 225 n (%) | 246,887 (6.8%) | 234,860 (7.0%) | 12,027 (4.5%) | 0.108 |
| | 2253 - n (%) | 371,949 (10.2%) | 319,731 (9.5%) | 52,218 (19.4%) | 0.285 |
| | 2254 - n (%) | 18,815 (0.5%) | 18,016 (0.5%) | 799 (0.3%) | 0.037 |
| | 2255 - n (%) | 41,455 (1.1%) | 40,826 (1.2%) | 629 (0.2%) | 0.116 |
| | 2256 - n (%) | 21,154 (0.6%) | 20,864 (0.6%) | 290 (0.1%) | 0.085 |
| | 2257 - n (%) | 22,990 (0.6%) | 22,621 (0.7%) | 369 (0.1%) | 0.084 |
| | 2258 - n (%) | 57,719 (1.6%) | 55,263 (1.6%) | 2,456 (0.9%) | 0.065 |
| | 2260 - n (%) | 157,395 (4.3%) | 151,377 (4.5%) | 6,018 (2.2%) | 0.125 |
| | 226 n (%) | 58,624 (1.6%) | 57,534 (1.7%) | 1,090 (0.4%) | 0.128 |
| | 2262 - n (%) | 45,934 (1.3%) | 43,859 (1.3%) | 2,075 (0.8%) | 0.052 |
| | 2263 - n (%) | 8,479 (0.2%) | 8,348 (0.2%) | 131 (0.0%) | 0.052 |
| | 2265 - n (%) | 134,215 (3.7%) | 125,166 (3.7%) | 9,049 (3.4%) | 0.019 |
| | 2266 - n (%) | 81,639 (2.2%) | 77,296 (2.3%) | 4,343 (1.6%) | 0.049 |
| | 2268 - n (%) | 100,882 (2.8%) | 90,266 (2.7%) | 10,616 (4.0%) | 0.071 |
| | 2270 - n (%) | 290,514 (8.0%) | 263,253 (7.8%) | 27,261 (10.2%) | 0.081 |
| | 2275 - n (%) | 52,650 (1.4%) | 50,459 (1.5%) | 2,191 (0.8%) | 0.064 |
| | 3895 - n (%) | 762,176 (21.0%) | 680,768 (20.2%) | 81,408 (30.3%) | 0.233 |
| ≥2 COVID vaccine doses | n (%) | 72,170 (2.0%) | 71,870 (2.1%) | 300 (0.1%) | 0.193 |
| 1-2 COVID vaccine doses | n (%) | 79,847 (2.2%) | 78,444 (2.3%) | 1,403 (0.5%) | 0.153 |
| 0 COVID vaccine doses | n (%) | 3,479,023 (95.8%) | 3,212,205 (95.5%) | 266,818 (99.4%) | 0.245 |
| Study period (bi-weekly, from Jan 2020) | 0 - n (%) | 10 (0.0%) | *5-9 | *1-5 | 0.001 |
| | 1 - n (%) | 13 (0.0%) | *8-12 | *1-5 | 0 |
| | 2 - n (%) | 101 (0.0%) | 101 (0.0%) | 0 (0.0%) | 0.147 |
| | 3 - n (%) | 131 (0.0%) | *126-130 | *1-5 | 0.079 |
| | 4 - n (%) | 1,048 (0.0%) | *1006-1010 | *38-42 | 0.162 |
| | 5 - n (%) | 12,254 (0.3%) | 10,887 (0.3%) | 1,367 (0.5%) | 0.048 |
| | 6 - n (%) | 16,312 (0.4%) | 12,596 (0.4%) | 3,716 (1.4%) | 0.04 |
| | 7 - n (%) | 23,678 (0.7%) | 18,963 (0.6%) | 4,715 (1.8%) | 0.09 |
| | 8 - n (%) | 29,917 (0.8%) | 25,799 (0.8%) | 4,118 (1.5%) | 0.059 |
| | 9 - n (%) | 30,957 (0.9%) | 27,856 (0.8%) | 3,101 (1.2%) | 0.061 |
| | 10 - n (%) | 58,964 (1.6%) | 55,379 (1.6%) | 3,585 (1.3%) | 0.008 |
| | 11- n (%) | 76,898 (2.1%) | 74,742 (2.2%) | 2,156 (0.8%) | 0.008 |

| | | | | | |
|---|---|---|---|---|---|
| | 12 - n (%) | 84,557 (2.3%) | 82,824 (2.5%) | 1,733 (0.6%) | 0.01 |
| | 13 - n (%) | 83,925 (2.3%) | 82,666 (2.5%) | 1,259 (0.5%) | 0.029 |
| | 14 - n (%) | 90,954 (2.5%) | 89,383 (2.7%) | 1,571 (0.6%) | 0.108 |
| | 15 - n (%) | 91,605 (2.5%) | 90,641 (2.7%) | 964 (0.4%) | 0.111 |
| | 16 - n (%) | 93,668 (2.6%) | 92,593 (2.8%) | 1,075 (0.4%) | 0.072 |
| | 17 - n (%) | 97,510 (2.7%) | 95,957 (2.9%) | 1,553 (0.6%) | 0.033 |
| | 18 - n (%) | 139,751 (3.8%) | 135,916 (4.0%) | 3,835 (1.4%) | 0.026 |
| | 19 - n (%) | 162,757 (4.5%) | 155,950 (4.6%) | 6,807 (2.5%) | 0.117 |
| | 20 - n (%) | 138,532 (3.8%) | 130,729 (3.9%) | 7,803 (2.9%) | 0.166 |
| | 21 - n (%) | 129,192 (3.6%) | 119,634 (3.6%) | 9,558 (3.6%) | 0.165 |
| | 22 - n (%) | 144,730 (4.0%) | 130,629 (3.9%) | 14,101 (5.3%) | 0.191 |
| | 23 - n (%) | 168,272 (4.6%) | 151,423 (4.5%) | 16,849 (6.3%) | 0.19 |
| | 24 - n (%) | 198,286 (5.5%) | 177,663 (5.3%) | 20,623 (7.7%) | 0.176 |
| | 25 - n (%) | 197,849 (5.4%) | 173,237 (5.2%) | 24,612 (9.2%) | 0.161 |
| | 26 - n (%) | 215,184 (5.9%) | 179,401 (5.3%) | 35,783 (13.3%) | 0.113 |
| | 27 - n (%) | 197,754 (5.4%) | 172,722 (5.1%) | 25,032 (9.3%) | 0.054 |
| | 28 - n (%) | 174,529 (4.8%) | 158,981 (4.7%) | 15,548 (5.8%) | 0 |
| | 29 - n (%) | 184,128 (5.1%) | 172,621 (5.1%) | 11,507 (4.3%) | 0.065 |
| | 30 - n (%) | 233,494 (6.4%) | 221,366 (6.6%) | 12,128 (4.5%) | 0.097 |
| | 31- n (%) | 274,403 (7.6%) | 257,838 (7.7%) | 16,565 (6.2%) | 0.156 |
| | 32 - n (%) | 279,677 (7.7%) | 262,863 (7.8%) | 16,814 (6.3%) | 0.277 |
| Prior mental health hospitalization | n (%) | 82,415 (2.3%) | 77,945 (2.3%) | 4,470 (1.7%) | 0.047 |
| Prior emergency department visit for mental health condition | n (%) | 200,301 (5.5%) | 188,832 (5.6%) | 11,469 (4.3%) | 0.062 |
| Prior outpatient mental health visit | n (%) | 796,358 (21.9%) | 747,646 (22.2%) | 48,712 (18.1%) | 0.102 |
| 1-year Baseline hospital-based costs | Mean (SD) | 1726.91 (10159.99) | 1764.50 (10206.32) | 1256.21 (9548.37) | 0.051 |
| | Median (Q1-Q3) | 0 (0-336) | 0 (0-336) | 0 (0-0) | 0.212 |
| 1-year Baseline emergency department costs | Mean (SD) | 312.49 (975.37) | 318.25 (971.10) | 240.44 (1024.54) | 0.078 |
| | Median (Q1-Q3) | 0 (0-278) | 0 (0-289) | 0 (0-143) | 0.114 |
| 1-yr Baseline medication costs | Mean (SD) | 610.82 (4140.93) | 623.54 (4226.39) | 451.54 (2857.37) | 0.048 |
| | Median (Q1-Q3) | 0 (0-50) | 0 (0-58) | 0 (0-14) | 0.152 |
| 1-yr Baseline outpatient-specialist costs | Mean (SD) | 885.05 (4744.63) | 902.28 (4790.92) | 669.21 (4115.09) | 0.052 |
| | Median (Q1-Q3) | 140 (0-642) | 145 (0-658) | 80 (0-457) | 0.161 |
| 1-yr Baseline outpatient-cancer costs | Mean (SD) | 341.02 (5058.17) | 357.13 (5174.60) | 139.28 (3259.14) | 0.05 |
| | Median (Q1-Q3) | 0 (0-0) | 0 (0-0) | 0 (0-0) | 0.073 |
| 1-yr Baseline outpatient-primary care costs | Mean (SD) | 380.35 (897.18) | 384.18 (913.27) | 332.40 (661.60) | 0.065 |
| | Median (Q1-Q3) | 247 (125-414) | 248 (128-416) | 222 (99-384) | 0.128 |
| 1-yr Baseline outpatient-lab costs | Mean (SD) | 53.37 (91.71) | 53.59 (92.17) | 50.53 (85.69) | 0.034 |
| | Median (Q1-Q3) | 21 (0-71) | 21 (0-71) | 21 (0-70) | 0.019 |
| 1-yr Baseline rehabilitation costs | Mean (SD) | 103.48 (1967.55) | 105.10 (1983.56) | 83.17 (1754.51) | 0.012 |
| | Median (Q1-Q3) | 0 (0-0) | 0 (0-0) | 0 (0-0) | 0.019 |
| 1-yr Baseline homecare costs | Mean (SD) | 247.50 (2241.14) | 249.45 (2226.99) | 223.09 (2411.07) | 0.011 |

| | | | | | |
|---|---|---|---|---|---|
| | Median (Q1-Q3) | 0 (0-0) | 0 (0-0) | 0 (0-0) | 0.077 |
| 1-yr Baseline mental health admission costs | Mean (SD) | 203.84 (5604.68) | 206.59 (5581.52) | 169.40 (5886.90) | 0.006 |
| | Median (Q1-Q3) | 0 (0-0) | 0 (0-0) | 0 (0-0) | 0.042 |
| 1-yr Baseline complex and continuing care costs | Mean (SD) | 139.15 (4539.02) | 140.46 (4587.99) | 122.74 (3873.60) | 0.004 |
| | Median (Q1-Q3) | 0 (0-0) | 0 (0-0) | 0 (0-0) | 0.003 |
| 1-yr Baseline long-term care costs | Mean (SD) | 0.08 (57.29) | 0.09 (59.51) | 0.02 (5.47) | 0.002 |
| | Median (Q1-Q3) | 0 (0-0) | 0 (0-0) | 0 (0-0) | 0.002 |

**Supplemental Figure E2:** Difference in annual median health care costs (95% confidence intervals in 2020 [illegible] starting 56 days after polymerase chain reaction test) for test-positive versus test-negative matched individu[illegible] and by sex

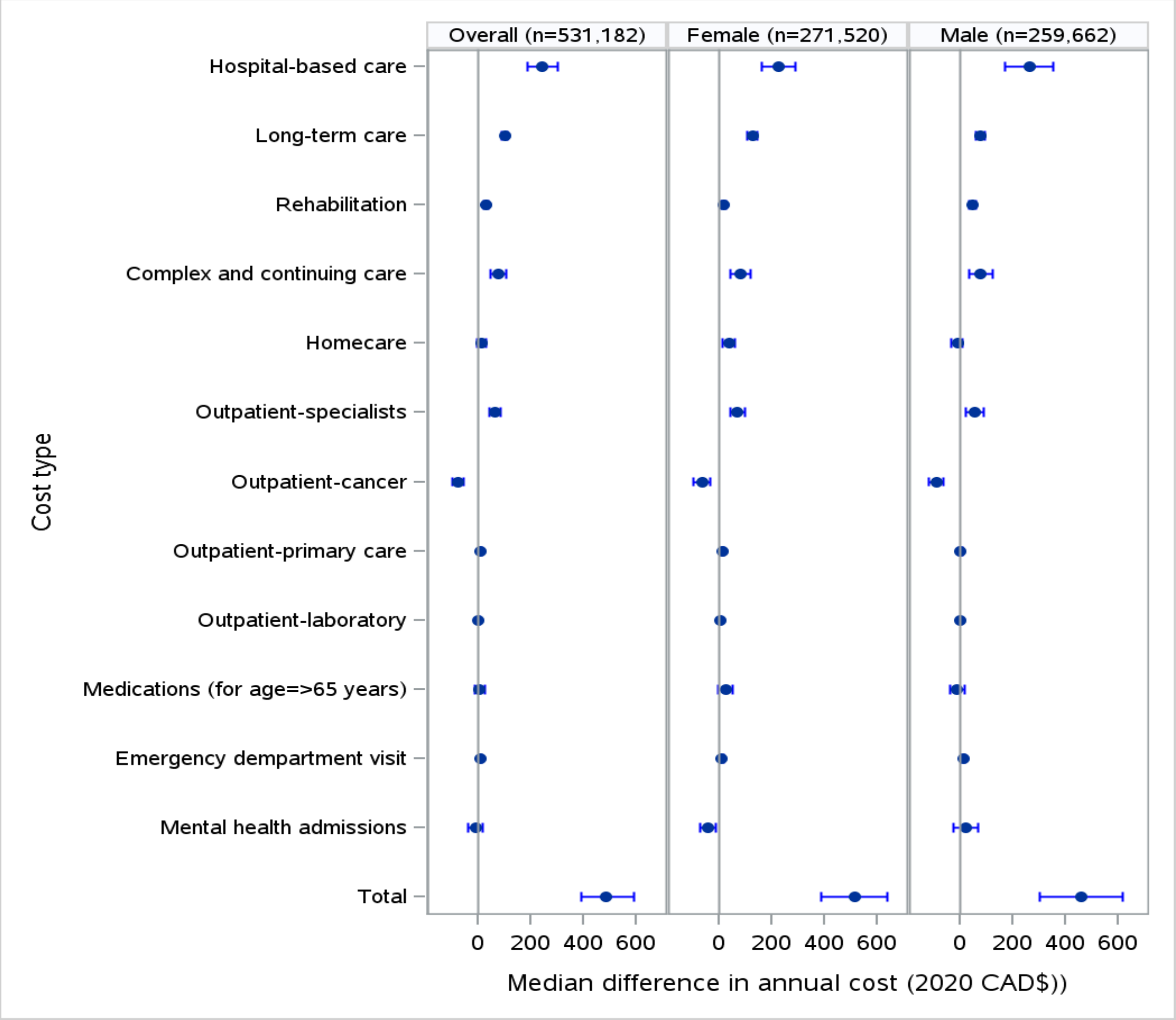

**Supplemental Figure E3:** Difference in annual 95th percentile health care costs (95% confidence intervals in 2020 $CAD, starting 56 days after polymerase chain reaction test) for test-positive versus test-negative matched individuals, overall and by sex

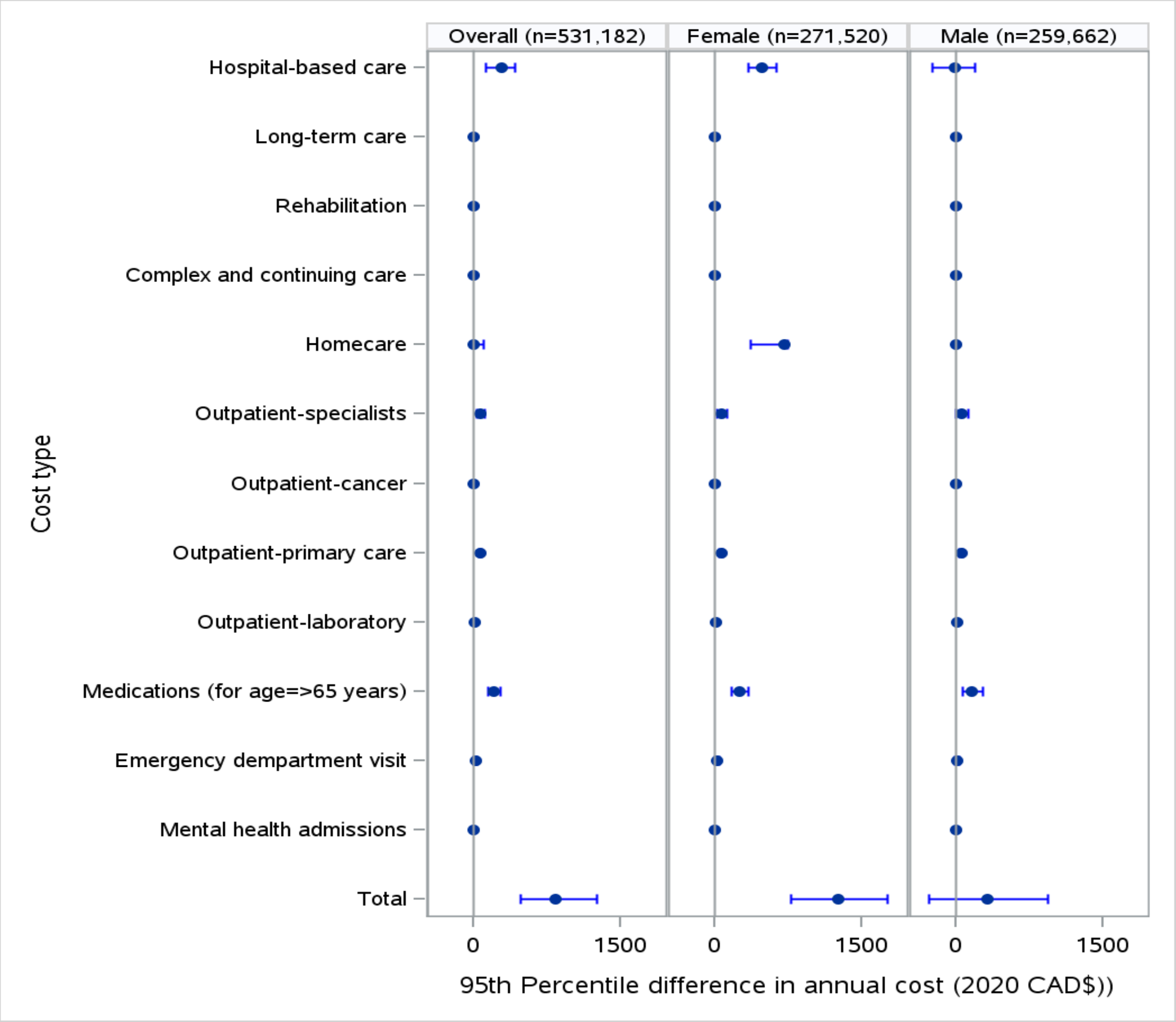

**Supplemental Figure E4**: Sensitivity analysis excluding long-term care costs

**(A)** Differences in annual mean health care costs (95% confidence intervals in 2020 $CAD, starting 56 days after polymerase chain reaction test) for test-positive versus test-negative matched individuals, overall and by sex

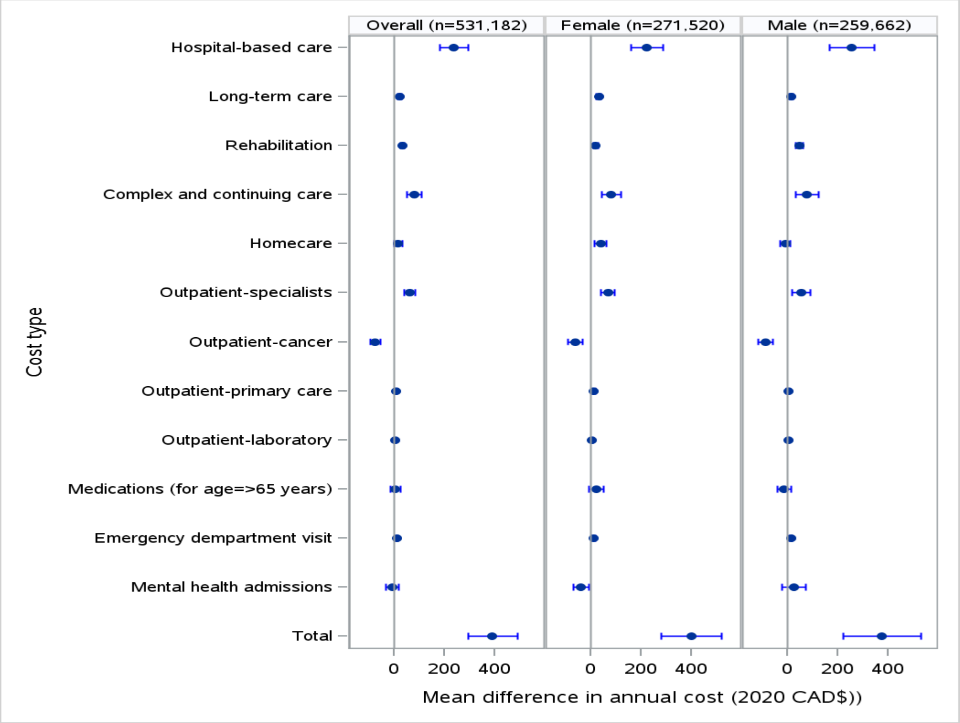

**(B)** Differences in annual median health care costs (95% confidence intervals in 2020 $CAD, starting 56 days after polymerase chain reaction test) for test-positive versus test-negative matched individuals, overall and by sex

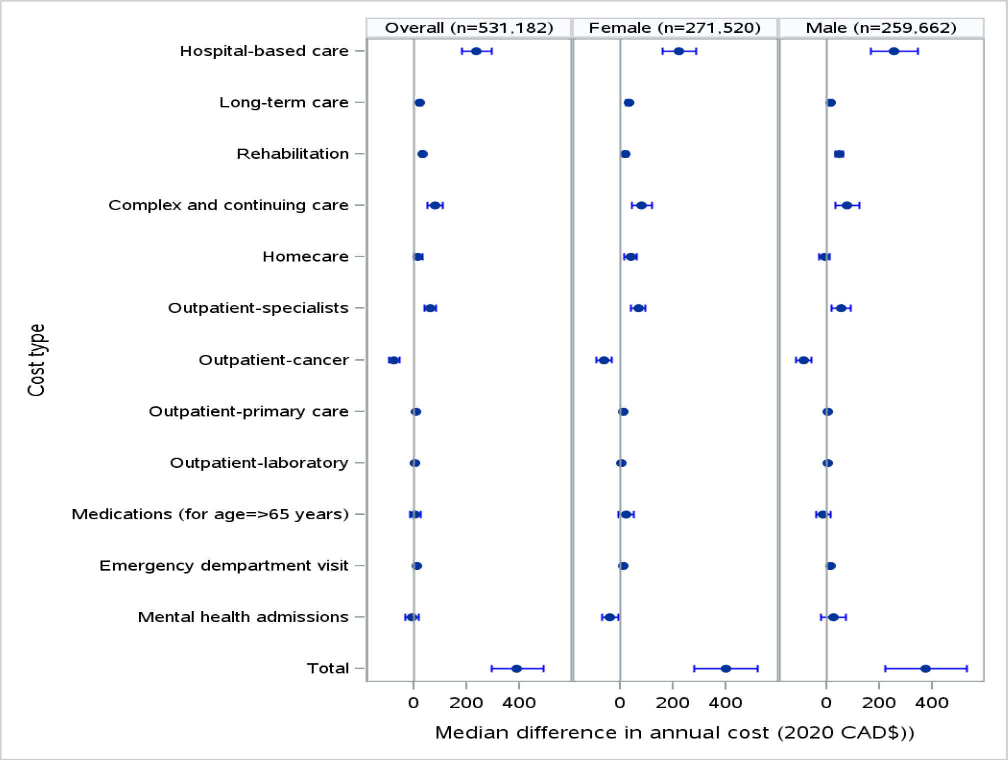

**(C)** Differences in annual 95th percentile health care costs (95% confidence intervals in 2020 $CAD, starting 56 days after polymerase chain reaction test) for test-positive versus test-negative matched individuals, overall and by sex

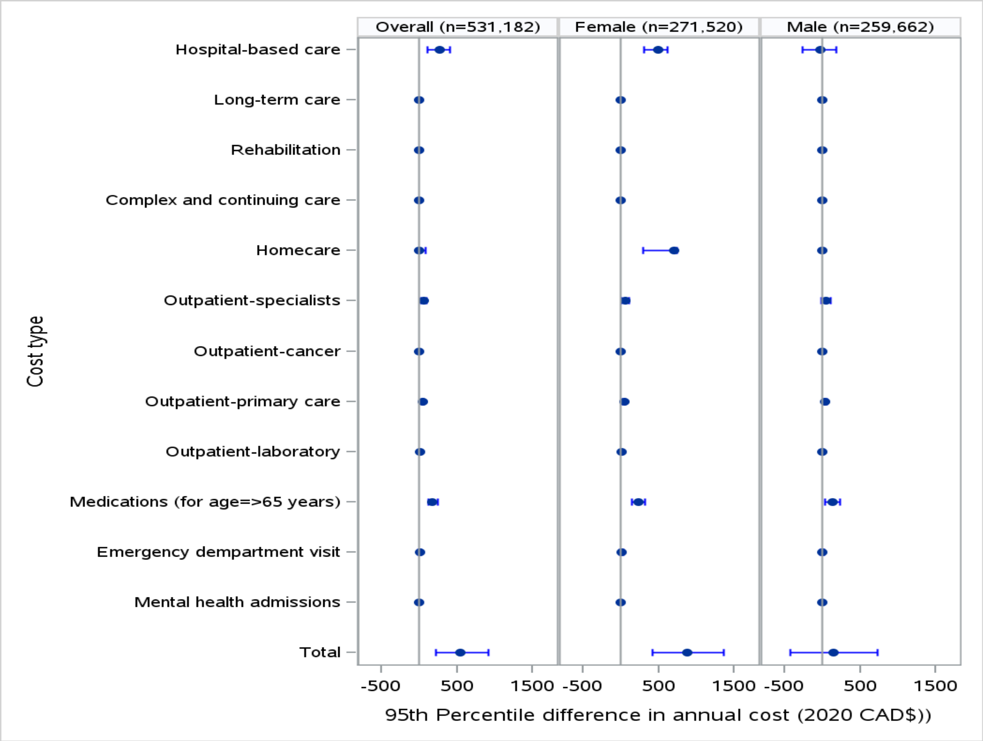

**(D)** Differences in annual 99th percentile health care costs (95% confidence intervals in 2020 $CAD, starting 56 days after polymerase chain reaction test) for test-positive versus test-negative matched individuals, overall and by sex

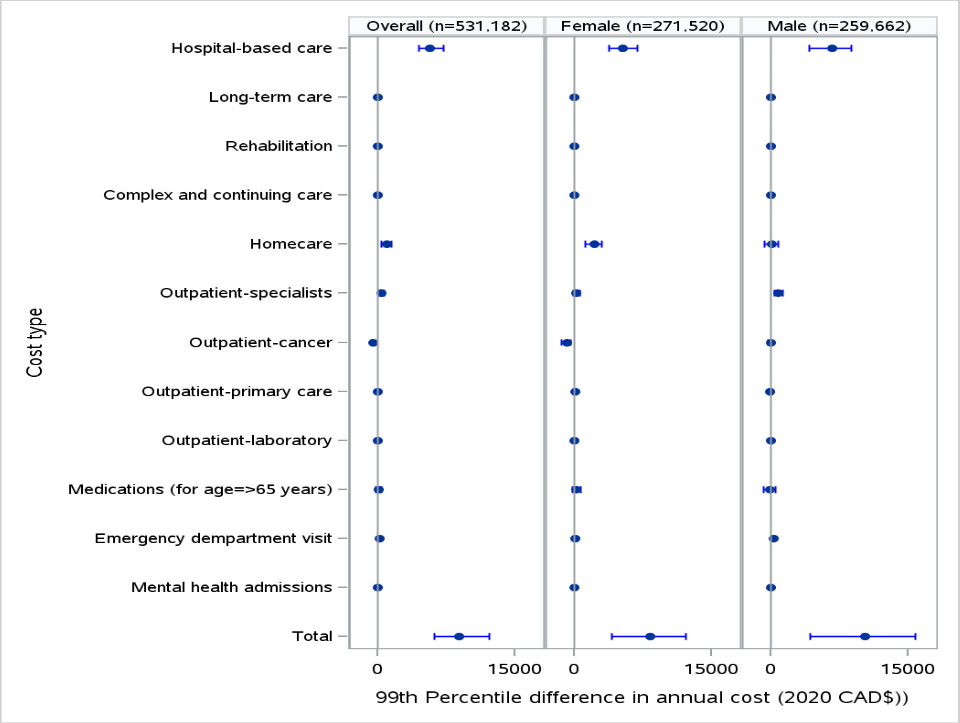

**Supplemental Figure E5**: Differences in 6-month health care costs (95% confidence intervals in 2020 $CAD, starting 56 days after polymerase chain reaction test) for test-positive versus test-negative matched individuals, overall and by sex, at the following distributions: (A) Mean, (B) Median, (C) 95th percentile, (D) 99th percentile

**(A) Mean**

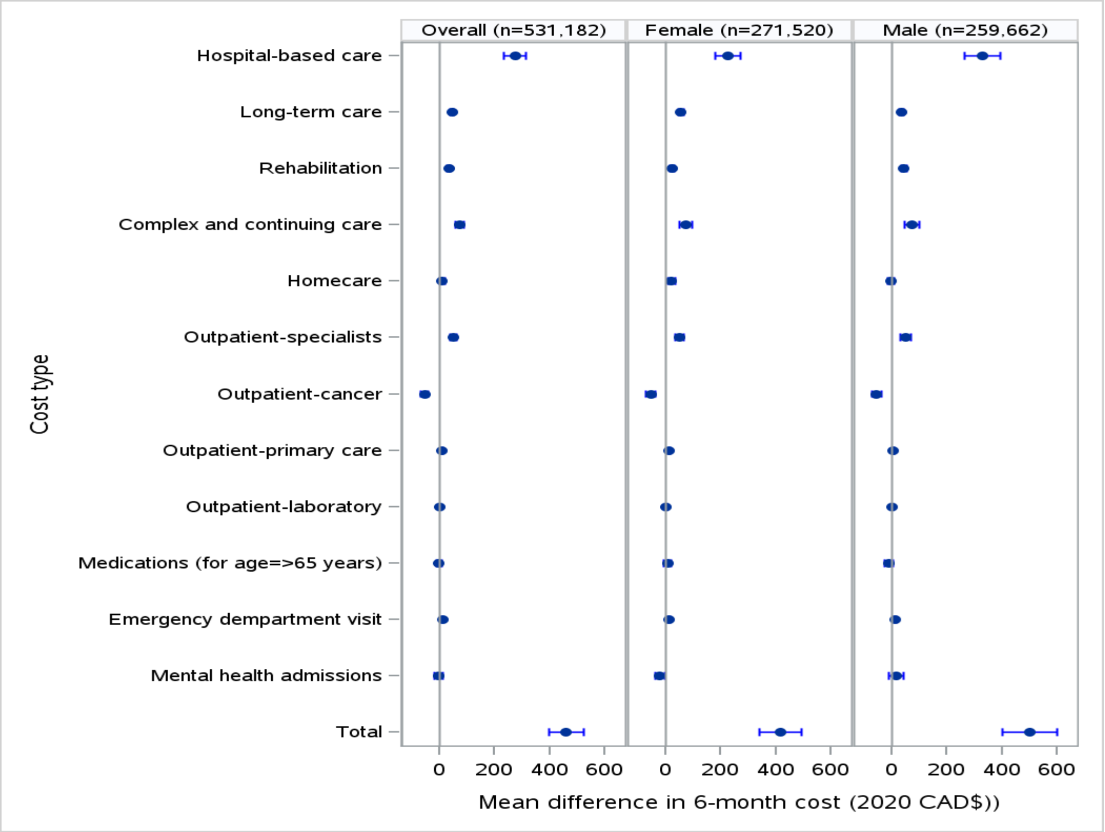

**(B) Median**

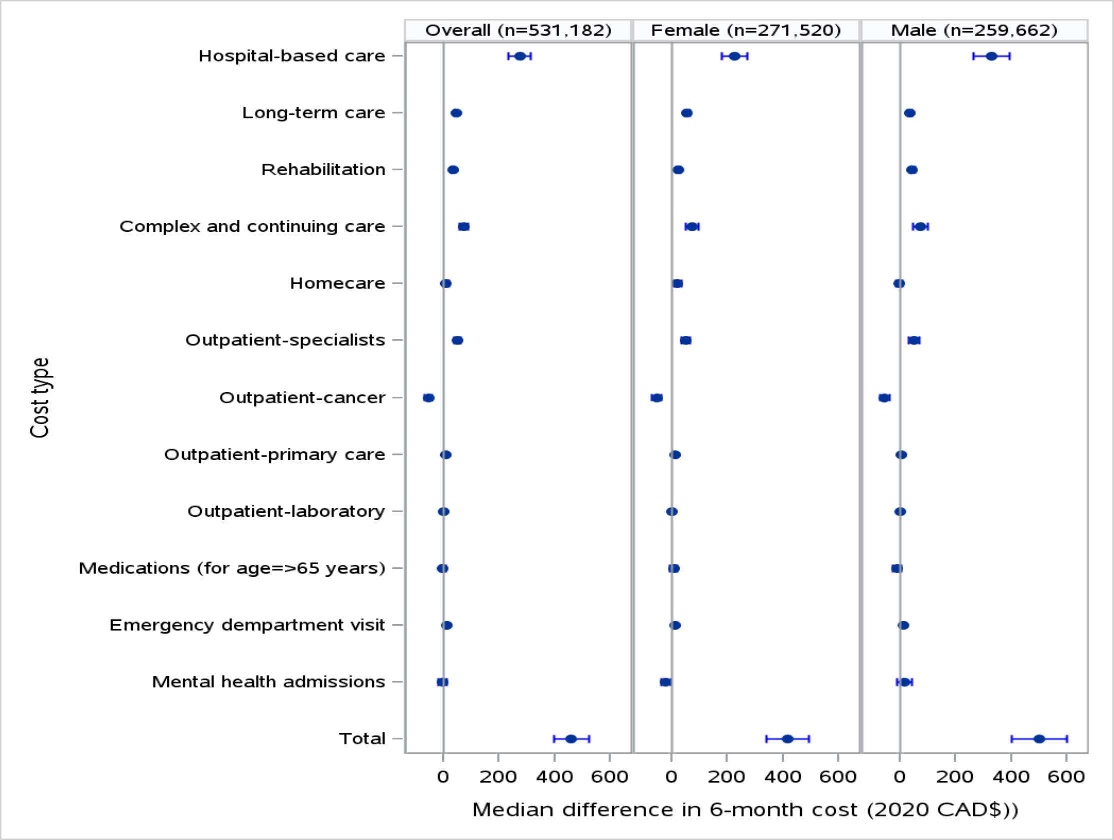

**(C) 95th percentile**

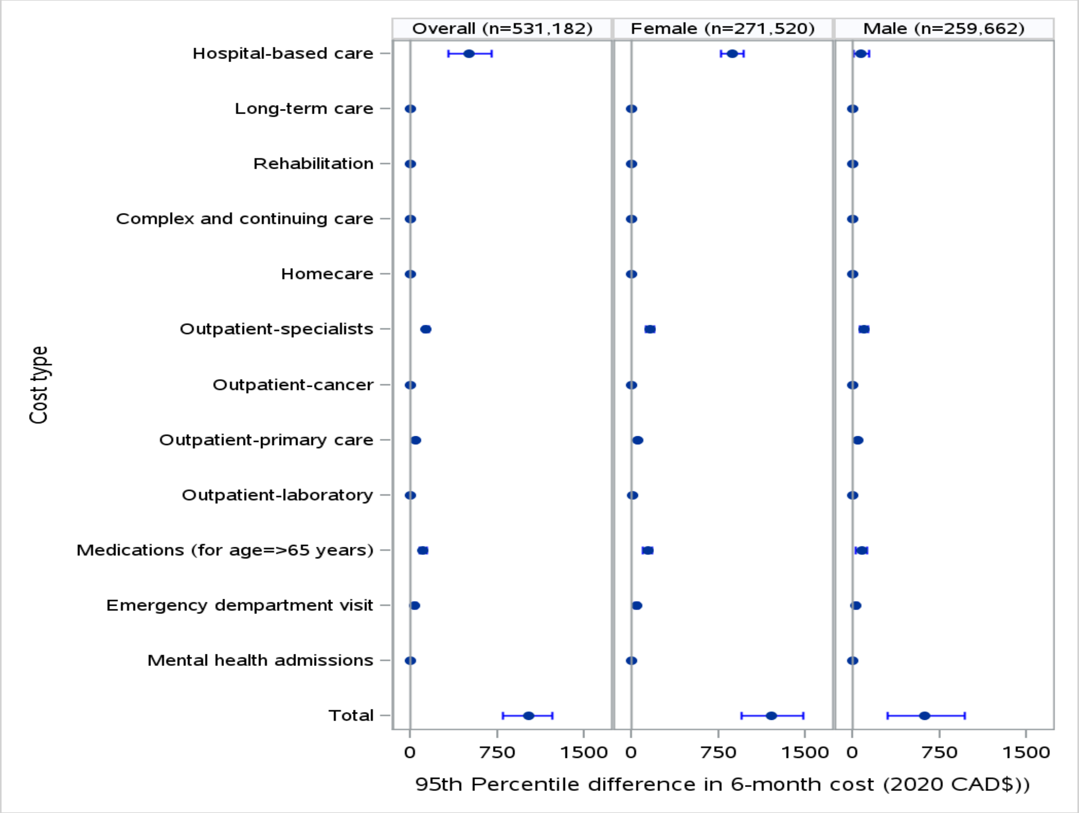

**(D) 99th percentile**

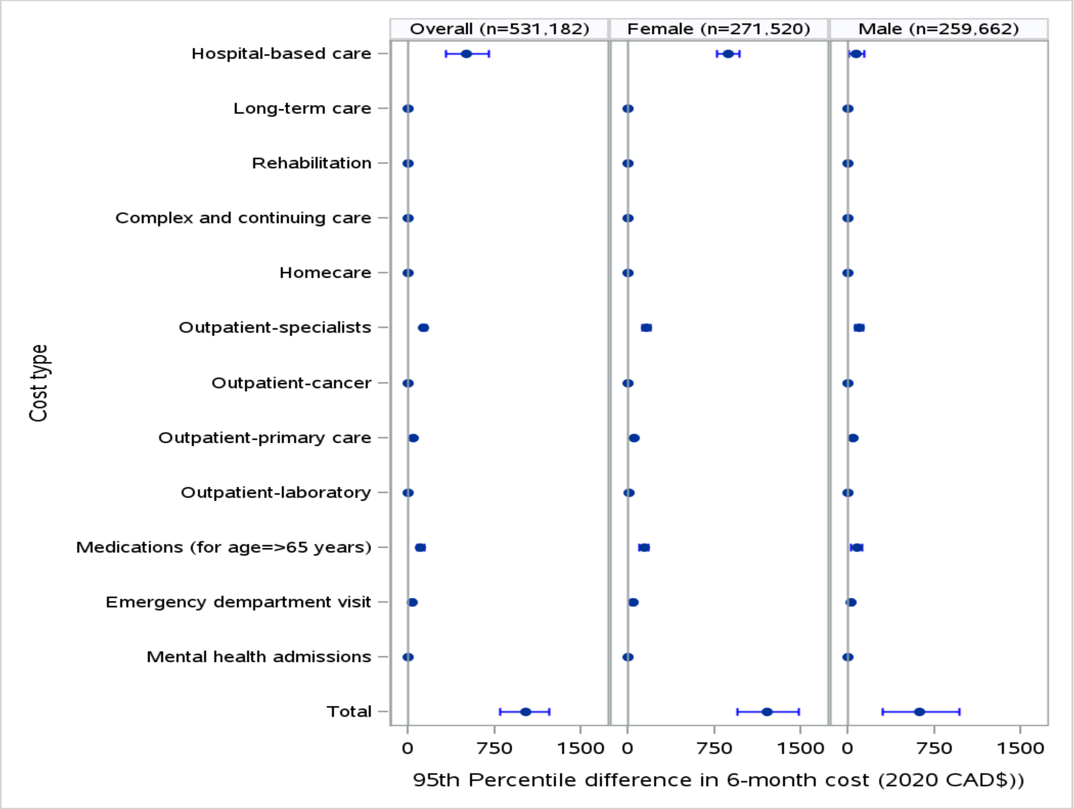

**Supplemental Table E5:** Risk differences between test-positive and test-negative people for being a high-cost user within 6 months and 1 year, overall and by sex, defined as (A) >95th percentile and (B) any post-acute health care cost ($>0)

| **(A) High-cost defined as cost>95th percentile** | | **6-months** | | | | | | **1-year** | | | | | |
|---|---|---|---|---|---|---|---|---|---|---|---|---|---|
| | **Outcome** | **# in T+** | **# in T-** | **∆% T+ vs. T-** | **RD lower 95%CI** | **RD upper 95%CI** | **P-value** | **# in T+** | **# in T-** | **∆% T+ vs. T-** | **RD lower 95%CI** | **RD upper 95%CI** | **P-value** |
| Overall (n=265,591) | Outpatient cancer | 13997 | 12564 | 0.0054 | 0.0042 | 0.0065 | <.0001 | 13630 | 12934 | 0.00 | 0.00 | 0.00 | <.0001 |
| | Rehabilitation | 14369 | 12726 | 0.0062 | 0.0050 | 0.0073 | <.0001 | 13575 | 12986 | 0.00 | 0.00 | 0.00 | 0.0002 |
| | Home care | 13756 | 12825 | 0.0035 | 0.0023 | 0.0047 | <.0001 | 13410 | 13161 | 0.00 | 0.00 | 0.00 | 0.1140 |
| | Mental health hospitalization | 14084 | 12482 | 0.0060 | 0.0049 | 0.0072 | <.0001 | 13548 | 13022 | 0.00 | 0.00 | 0.00 | 0.0008 |
| | Complex continuing care | 14418 | 12183 | 0.0084 | 0.0073 | 0.0096 | <.0001 | 13998 | 12595 | 0.01 | 0.00 | 0.01 | <.0001 |
| | Long-term care | 13794 | 12943 | 0.0032 | 0.0020 | 0.0044 | <.0001 | 13687 | 13060 | 0.00 | 0.00 | 0.00 | <.0001 |
| Female (n=135,760) | Outpatient cancer | 7622 | 6613 | 0.0074 | 0.0058 | 0.0091 | <.0001 | 7073 | 6502 | 0.00 | 0.00 | 0.01 | <.0001 |
| | Rehabilitation | 8353 | 7108 | 0.0092 | 0.0075 | 0.0109 | <.0001 | 7591 | 6976 | 0.00 | 0.00 | 0.01 | <.0001 |
| | Home care | 7530 | 6984 | 0.0040 | 0.0023 | 0.0057 | <.0001 | 7380 | 7202 | 0.00 | 0.00 | 0.00 | 0.1275 |
| | Mental health hospitalization | 8269 | 7179 | 0.0080 | 0.0063 | 0.0097 | <.0001 | 7890 | 7584 | 0.00 | 0.00 | 0.00 | 0.0104 |
| | Complex continuing care | 8618 | 7218 | 0.0103 | 0.0086 | 0.0121 | <.0001 | 8384 | 7477 | 0.01 | 0.00 | 0.01 | <.0001 |
| | Long-term care | 9185 | 8463 | 0.0053 | 0.0035 | 0.0072 | <.0001 | 8988 | 8394 | 0.00 | 0.00 | 0.01 | <.0001 |
| Male (n=129,831) | Outpatient cancer | 6375 | 5951 | 0.0033 | 0.0017 | 0.0049 | <.0001 | 6557 | 6432 | 0.00 | 0.00 | 0.00 | 0.2495 |
| | Rehabilitation | 6016 | 5618 | 0.0031 | 0.0015 | 0.0046 | 0.0001 | 5984 | 6010 | 0.00 | 0.00 | 0.00 | 0.8055 |
| | Home care | 6226 | 5841 | 0.0030 | 0.0014 | 0.0046 | 0.0003 | 6030 | 5959 | 0.00 | 0.00 | 0.00 | 0.5019 |
| | Mental health hospitalization | 5815 | 5303 | 0.0039 | 0.0024 | 0.0055 | <.0001 | 5658 | 5438 | 0.00 | 0.00 | 0.00 | 0.0305 |
| | Complex continuing care | 5800 | 4965 | 0.0064 | 0.0049 | 0.0079 | <.0001 | 5614 | 5118 | 0.00 | 0.00 | 0.01 | <.0001 |
| | Long-term care | 4609 | 4480 | 0.0010 | -0.0004 | 0.0024 | 0.1671 | 4699 | 4666 | 0.00 | 0.00 | 0.00 | 0.7269 |

| (B) Any post-acute health care cost | | 6-months | | | | | | 1-year | | | | | |
|---|---|---|---|---|---|---|---|---|---|---|---|---|---|
| | **Outcome** | **# in T+** | **# in T-** | **∆% T+ vs. T-** | **RD lower 95%CI** | **RD upper 95%CI** | **P-value** | **# in T+** | **# in T-** | **∆% T+ vs. T-** | **RD lower 95%CI** | **RD upper 95%CI** | **P-value** |
| Overall (n=265,591) | Outpatient cancer | 1739 | 2369 | -0.0024 | -0.0028 | -0.0019 | <.0001 | 2428 | 3071 | 0.00 | 0.00 | 0.00 | <.0001 |
| | Rehabilitation | 744 | 308 | 0.0016 | 0.0014 | 0.0019 | <.0001 | 954 | 570 | 0.00 | 0.00 | 0.00 | <.0001 |
| | Home care | 10740 | 9780 | 0.0036 | 0.0026 | 0.0046 | <.0001 | 13109 | 12339 | 0.00 | 0.00 | 0.00 | <.0001 |
| | Mental health hospitalization | 698 | 717 | -0.0001 | -0.0003 | 0.0002 | 0.6127 | 1008 | 1127 | 0.00 | 0.00 | 0.00 | 0.0097 |
| | Complex continuing care | 1138 | 554 | 0.0022 | 0.0019 | 0.0025 | <.0001 | 1327 | 792 | 0.00 | 0.00 | 0.00 | <.0001 |
| | Long-term care | 1064 | 432 | 0.0024 | 0.0021 | 0.0027 | <.0001 | 1390 | 682 | 0.00 | 0.00 | 0.00 | <.0001 |
| Female (n=135,760) | Outpatient cancer | 1054 | 1385 | -0.0024 | -0.0031 | -0.0017 | <.0001 | 1468 | 1788 | 0.00 | 0.00 | 0.00 | <.0001 |
| | Rehabilitation | 301 | 136 | 0.0012 | 0.0009 | 0.0015 | <.0001 | 404 | 280 | 0.00 | 0.00 | 0.00 | <.0001 |
| | Home care | 5894 | 5214 | 0.0050 | 0.0035 | 0.0065 | <.0001 | 7165 | 6545 | 0.00 | 0.00 | 0.01 | <.0001 |
| | Mental health hospitalization | 299 | 336 | -0.0003 | -0.0006 | 0.0001 | 0.1408 | 447 | 531 | 0.00 | 0.00 | 0.00 | 0.0070 |
| | Complex continuing care | 562 | 257 | 0.0022 | 0.0018 | 0.0027 | <.0001 | 667 | 367 | 0.00 | 0.00 | 0.00 | <.0001 |
| | Long-term care | 619 | 228 | 0.0029 | 0.0025 | 0.0033 | <.0001 | 807 | 371 | 0.00 | 0.00 | 0.00 | <.0001 |
| Male (n=129,831) | Outpatient cancer | 685 | 984 | -0.0023 | -0.0029 | -0.0017 | <.0001 | 960 | 1283 | 0.00 | 0.00 | 0.00 | <.0001 |
| | Rehabilitation | 443 | 172 | 0.0021 | 0.0017 | 0.0025 | <.0001 | 550 | 290 | 0.00 | 0.00 | 0.00 | <.0001 |
| | Home care | 4846 | 4566 | 0.0022 | 0.0007 | 0.0036 | 0.0028 | 5944 | 5794 | 0.00 | 0.00 | 0.00 | 0.1492 |
| | Mental health hospitalization | 399 | 381 | 0.0001 | -0.0003 | 0.0006 | 0.5187 | 561 | 596 | 0.00 | 0.00 | 0.00 | 0.3010 |
| | Complex continuing care | 576 | 297 | 0.0021 | 0.0017 | 0.0026 | <.0001 | 660 | 425 | 0.00 | 0.00 | 0.00 | <.0001 |
| | Long-term care | 445 | 204 | 0.0019 | 0.0015 | 0.0022 | <.0001 | 583 | 311 | 0.00 | 0.00 | 0.00 | <.0001 |
